# Neural signatures of risk-taking adaptions across health, bipolar disorder, and lithium treatment

**DOI:** 10.1101/2023.03.13.23287200

**Authors:** Jacqueline Scholl, Priyanka Panchal, Natalie Nelissen, Lauren Z Atkinson, Nils Kolling, Kate EA Saunders, John Geddes, Matthew FS Rushworth, Anna C Nobre, Paul J Harrison, Catherine J Harmer

**Author notes:** Corresponding author, additional contact information, 0033663543810.

## Abstract

Cognitive and neural mechanisms underlying bipolar disorder (BD) and its treatment are still poorly understood. Here we examined the role of adaptations in risk-taking using a reward-guided decision-making task.

We recruited volunteers with high (n=40) scores on the Mood Disorder Questionnaire, MDQ, suspected of high risk for bipolar disorder and those with low-risk scores (n=37). We also recruited patients diagnosed with BD who were assigned (randomized, double-blind) to six weeks of lithium (n=19) or placebo (n=16) after a two-week baseline period (n=22 for FMRI). Participants completed mood ratings daily over 50 (MDQ study) or 42 (BD study) days, as well as a risky decision-making task and functional magnetic resonance imaging. The task measured adaptation of risk taking to past outcomes (increased risk aversion after a previous win vs. loss, ‘outcome history’).

While the low MDQ group was risk averse after a win, this was less evident in the high MDQ group and least so in the patients with BD. During fMRI, ‘outcome history’ was linked to medial frontal pole activation at the time of the decision and this activation was reduced in the high risk MDQ vs. the low risk MDQ group. While lithium did not reverse the pattern of BD in the task, nor changed clinical symptoms of mania or depression, it changed reward processing in the dorsolateral prefrontal cortex.

Participants’ modulation of risk-taking in response to reward outcomes was reduced as a function of risk for BD and diagnosed BD. These results provide a model for how reward may prime escalation of risk-related behaviours in bipolar disorder and how mood stabilising treatments may work.

**Key points:** **Question:** Do bipolar disorder and lithium treatment change adaptation of risk-taking over time?

**Findings:** Across an observational study and a randomized controlled trial, we found that while participants modulate their risk taking in a gambling task over time, this was reduced as a function of risk for bipolar disorder. Neurally, this was accompanied by changes in reward memory traces in medial frontal pole.

**Meaning:** The results show that bipolar disorder is linked to a reduction in adaptation of risk- taking to the environment, suggesting a possible computational mechanism and treatment target.

## Introduction

Bipolar disorder (BD) is typically characterized by episodes of depression or mania, lasting weeks and months. Lithium is the most effective mood stabiliser for management of BD, reducing the frequency of both manic and depressive episodes (1). While fluctuating mood episodes have traditionally be seen as lasting weeks or months, more recent work has shown that, in fact, patients with BD show large day-to-day fluctuations in mood even when symptoms are in the non-clinical range (2) and that this is affected by lithium treatment (3). Understanding the processes underpinning bipolar disorder may help us develop and assess more effective treatment approaches.

From a computational psychiatry perspective, two causes for mood fluctuations in bipolar disorder could be considered. First, mood fluctuations could be the result of either increased and prolonged responses to valanced outcomes. Recent work from the field of reinforcement learning has suggested that destabilizing positive feedback cycles between mood and perceptions of rewards may contribute to BD (4–7): In people with subclinical symptoms of BD, positive or negative surprises were found to affect the neural and behavioural responses to reward and punishments. In particular, symptoms were associated with an increase in reward value after a positive surprise. This kind of reward sensitivity has been linked to later changes in mood, suggesting a route by which escalation of reward responses may translate into clinical symptoms (4). Second, mood fluctuations could be the result of reduced behaviours that stabilize mood. Using momentary ecological monitoring has revealed that in the healthy state, when mood fluctuates, people self-report using strategies to re-establish mood homeostasis such as engaging with aversive activities when they are in a good mood

(8). This strategy is reduced in people with depression or low mood (9). However, it is yet unclear whether regulating behaviour is also reduced in BD. In the lab, adaptations of behaviour to past outcomes have been studied in the field of decision-making, revealing temporal interdependencies. For example, people show ‘biases’ such as ‘loss chasing’ (10) (taking more risks to try and recover losses). Here, we used a lab-based task that allowed us to test the impact of BD and its’ treatment on both putative processes.

Optimal decision making involves interplay between frontostriatal systems, which play a role in motivation, reward value and its regulation. The ventral striatum and the ventromedial prefrontal cortex (vmPFC) are implicated in reward anticipation as well as its hedonic impact (11,12). vmPFC is further implicated in the evaluation of options (13), including tracking of past reward outcomes (14). We would therefore expect that if bipolar disorder affects the adaptation of behaviour to past outcomes, these signals in the vmPFC should be changed. By contrast, activity in the dorsolateral PFC is associated with regulation of behaviour towards reward, including self-regulation of reward craving (15,16). Previous work has linked bipolar disorder to increased reward related striatal signalling, coupled with altered patterns of ventromedial and dorsolateral PFC engagement (17) and interaction (18), while a meta-analysis (19) has highlighted a role for orbitofrontal cortex abutting dlPFC.

Here, we have built on these findings to test whether a gradient across a bipolar disorder spectrum (i.e. from low risk to diagnosed bipolar disorder), was linked to changed behavioural adaptation (risk taking) from trial to trial in response to reward/loss outcomes. For this, we recruited 40 volunteers with high scores on the mood disorder questionnaire (MDQ (20)), at suspected high risk for bipolar disorder, 37 volunteers with low scores, and 35 treatment seeking patients with diagnosed BD (n=22 for FMRI). To assess whether behaviour and naturally occurring daily-life mood fluctuations were related, participants completed up to 50 longitudinal testing sessions at home. To understand the neural mechanisms of risk adaptation behaviour, we measured brain activity with fMRI. To test the causal effect of a commonly prescribed mood-stabilizing drug, lithium, 19 patients were randomly assigned to receive six weeks of lithium treatment (dose titrated individually to plasma levels of 0.6-1 mmol/L) and 16 to placebo treatment in a double-blind design.

We hypothesized that BD and risk for BD would be associated with reduced adaption of risk taking behaviour (i.e. choice being less connected to previous experience of a win or a loss), which would be associated with changes in vmPFC and dlPFC signalling of previous win/loss experiences during fMRI. We also hypothesized that these behavioural and neuroimaging differences would be normalised following six weeks of lithium vs placebo treatment in BD.

## METHODS

### Participants

Participants were recruited in two separate studies (see below). The non-interventional study was approved by the local ethics committee (MSD-IDREC-C2-2014-023) and the interventional study by the National Research Ethics Service Committee South Central – Oxford A (15/SC/0109) and the Oxford Health NHS Foundation Trust. Participants gave informed consent and were reimbursed for taking part in the study.

#### Volunteers at suspected high vs low risk of bipolar disorder

Participants were recruited through local advertisement and from pools of previous participants. In an online pre-screening session, participants completed the Mood Disorders Questionnaire (MDQ (20)), a self-report screening instrument to identify risk for bipolar disorder. Participants were only invited for a full screening session if they scored either <5 points (‘low MDQ’ group, n=37 included, at presumed low risk for bipolar disorder); or ≥ 7 (‘high MDQ’ group, n=40 included). The screening verified that several of these symptoms measured with the MDQ happened during the same period of time. Structured clinical interviews with the SCID revealed that 5 of this group met criteria for bipolar disorder, despite not having received a formal diagnosis or seeking treatment. See supplementary method [1A] for detailed exclusion criteria.

#### Patients with BD

Participants were recruited through the BD Research Clinic (Oxford). All participants met criteria for BD-I (n=7), BD-II (n=27) or BD not otherwise specified (BD-NOS, n=1), based on structured clinical interview. All participants were outside major mood episodes requiring immediate treatment. Full exclusion criteria are provided in the supplementary materials [1B]. Participants were assigned to placebo (n=16) or lithium (n=19), in a randomised double-blind design, see below.

### Study design

#### Volunteers

We measured participants’ mood and behaviour in a cognitive task longitudinally five times a week over ten weeks. Brain activity during the same task was measured during an MRI scan. The data here were part of a larger study (supplementary method [1B]).

#### Patients with BD

This study was a randomised, 6-week, double-blind, placebo-controlled trial (21). See supplementary method [1B] for full information. All participants underwent a two-week pre-randomization phase (‘baseline’) during which they completed the cognitive task and mood ratings daily at home. Due to logistic challenges, for some participants this phase lasted longer than two weeks. For the next phase (6 weeks), participants were pseudo-randomly assigned to receive either lithium (starting dose of 400mg and then titrated to plasma levels of 0.6-1 mmol/L) or placebo in a double-blind design. Only 22 participants were fMRI compatible. Participants were invited to complete online weekly assessments of depression symptoms with the Quick Inventory of Depressive Symptomatology (QIDS, (22)) and symptoms of Mania with the Altman Self Rating Mania Scale (23).

Throughout, we performed two types of group comparisons. First, we compared across risk of BD (i.e. group as ordered factor (24) in regressions, Low MDQ ≤ High MDQ ≤ patients with BD), subsequently referred to as ‘bipolar disorder gradient’. Ordered factors in regression imply a relationship of order between the groups, this does not have to be a linear relationship (i.e. the difference low MDQ to high MDQ can be larger or smaller [but of same sign] than the difference high MDQ to patients with BD). MDQ was not measured in the patient group. As this involved data from the BD group before assignment to lithium or placebo, all participants were included. Significant results were post-hoc followed up comparisons of the individual groups (t-tests). Second, we tested for the effects of lithium treatment as drug (lithium/placebo) x time point (pre, i.e. baseline/post) interactions.

### ‘Wheel of fortune’ task

#### Trial structure

On each trial of the task, participants were given two options shown side-by-side. In the at-home version, these were wheels of fortune (Figure 1A). In the fMRI version, they were instead presented as bars. Each option had three attributes: probability of winning vs. losing (size of green vs. red area), magnitude of possible gain (number on green area, 10 to 200), and magnitude of possible loss (number on red area, also 10 to 200). After participants chose one option, the wheel of fortune started spinning and then randomly landed on either win or loss. Finally, participants were shown their updated total score. The experiment was designed so that most choices were difficult, i.e., the options were very similar in expected value, i.e. relative utility (reward magnitude * probability; 90% of choices were not more than 20 points apart; 76% not more than 5 points apart, Figure 1B, Figure S1).

**Figure 1.**
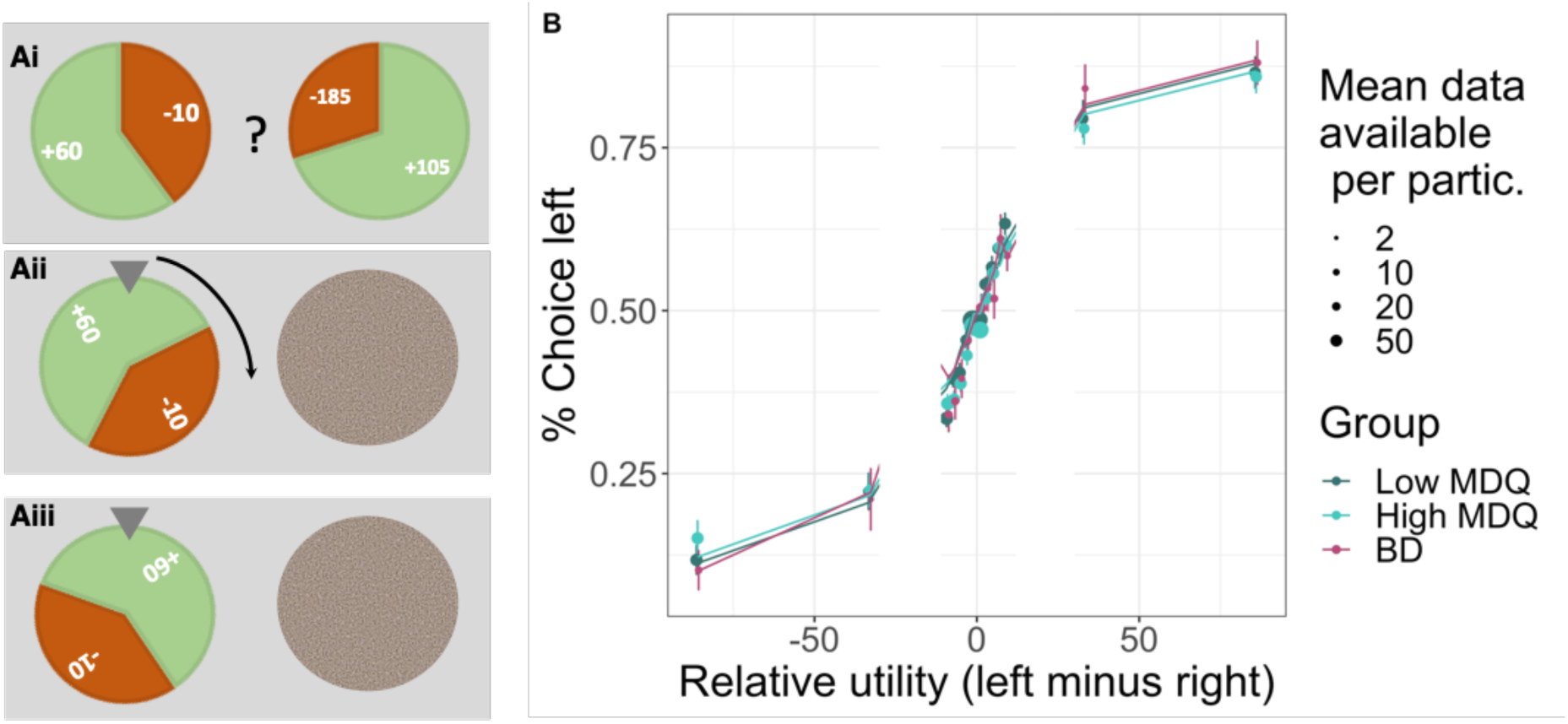
Task design and longitudinal behaviour. A) On each trial, participants chose between two gambles (‘wheel of fortune’) that differed in their probability of winning or losing points and in the number of points that could be won or lost. Once participants had chosen an option, the alternative was hidden, and the chosen wheel started spinning until finally landing on the win or loss. B) Participants’ choices (left vs. right option) were guided by the relative utilities (reward utility – i.e., probability * magnitude – minus loss utility): the higher the utility of the left option, the more it was chosen. The computational model (lines) captured behaviour (dots with error bars) well. Data were combined across all testing sessions (up to 50) per participant (20 trials per session). Error bars show the standard error of the mean, and the size of the dots indicates the number of data points available.

#### Timings and number of trials

Each day, participants rated their positive and negative mood using the Positive and Negative Affect Schedule – Short Form, PANAS-SF (25). They also gave an overall rating of their mood (‘How are you feeling’, referred to here as ‘happiness VAS’) using a slider ranging from ‘very unhappy’ (red sad face drawing) to ‘very happy’ (green smiley face). They then played 20 trials of the task. After the task, they repeated the happiness VAS.

In the fMRI scanner, participants played 100 trials. All timings were jittered. From the onset of options until participants could make a choice: 1-2 sec; delay between participants’ response and outcomes shown: 2.7 to 7.7 sec; duration of outcome shown: 1-3 sec; duration of total score shown: 1-9 sec; ITI: 1-9 sec.

### Behaviour

Behavioural data were analysed in R (26) (version 4.0.2) and Matlab. R-packages: Stan (27), BRMS (28,29), dplyr (30), ggpubr (31), sjPlot (32), compareGroups (33), emmeans (34), ggsci (35).

#### Group comparisons

To compare groups, instead of a standard ANOVA procedure which tests for any differences between groups, we tested for a systematic effect, i.e. bipolar disorder gradient (group as ordered factor (24), Low MDQ ≤ High MDQ ≤ patients with BD) in linear regressions, also controlling for age and gender. Models used the BRMS toolbox interface for Stan (supplementary methods 2). For this and all subsequent analyses, we used Bayesian Credible Intervals (36) to establish significance by the 95% CI not including zero.

### Computational models

#### Decision making

We used a computational model to capture participants’ choices. The model first computed the overall expected (‘utility’) of each option, then made a choice (left or right option) depending on which option had the better utility, but also allowing for some random choice behaviour (37,38).

First, the model compared the options’ utilities as displayed at the time of choice on the current trial, i.e., probability (prob) x magnitude (mag). We allowed for individual differences in sensitivity to the loss vs. reward utility (λ). We also included in the model a measure of adaptions of risk taking (i.e. loss vs win sensitivity) to past outcomes (‘outcome history’). Specifically, a parameter (γ) changed the weighting of the loss utility on the current trial depending on whether the previous trial’s outcome was a win or a loss (i.e., γ>0 means increased sensitivity to losses after a win on the previous trial).

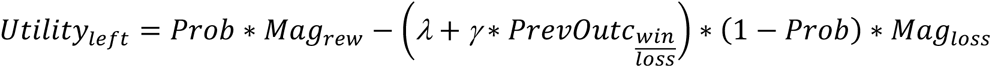

To decide which option to choose, the model compared the utilities of the left and right options taking into account each participant’s ‘randomness’ (inverse temperature (β), higher numbers indicating higher choice consistency):

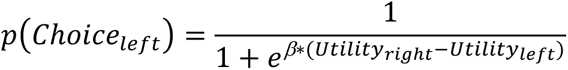

To allow fitting of individual sessions (20 trials), a Bayesian approach was implemented that allowed specifying priors for each parameter (supplementary methods [2C]). The model was validated using simulations and model comparisons (Table S1-3 and supplementary methods [2A-B]).

#### Group differences

To assess group differences, we entered the session-wise parameters into hierarchical regressions (using BRMS). This allowed us to take into account that parameters might change over days of testing, as well as individual differences in the means and variability (standard deviation) across sessions. For example:

Mean: invTemp(β) ∼ 1 + day + group+ Age + Gender + (1 + day | ID),

And error term: sigma ∼ 1 + group + Age + Gender + (1|ID))

The effect of lithium (vs. placebo) was tested analogously:

Mean: invTemp(β) ∼ 1 + day + group*pre, i.e. baseline/post+ Age + Gender + number_days_baseline + (1 + day | ID)

These models were used for group comparisons of mean parameters (supplementary methods 2C+D). Variabilities of parameters over days were not compared as model validation (Table S1) suggested poor recovery. Mood data (positive and negative PANAS, happiness VAS) were analysed using similar regressions (supplementary methods 2D) to assess group differences in mood (mean or variability) or the relationship between task outcomes and changes in happiness VAS.

### Model-free analyses of behaviour

To test that participants could perform the task, i.e., that their choices were sensitive to expected value, we binned their choices (% left vs. right option) according to the overall utility difference between the two options (i.e., left vs. right reward utility minus loss utility, utility=probability*magnitude).

To test sensitivity to risk of losses, as has been previously reported to be affected in BD (39,40), we refined the binning of choices (as above) by further splitting the data according to win and loss utility (i.e. probability * magnitude).

We next analysed behaviour for adaptions of risk taking to past outcomes by considering how participants change their behaviour – here risk-taking (avoidance of potential losses) – based on win/loss outcomes on previous trials (‘outcome history’ effect). For this, we computed how their choices differed after a win or loss on the previous trial (difference % choosing option with lower potential loss [loss utility] after win minus after a loss). We focused on the most extreme (lowest/highest) loss utility bins from the analyses above (‘taking loss utility more into account’) as adaptation to past trial outcomes by taking loss more into account (i.e. a multiplicative effect) should most strongly affect choices the more dissimilar the loss utilities of the two options.

### MRI acquisition

Data from all 77 high and low MDQ volunteers and 13 patients with BD were collected on a 3T Siemens Magneton Trio. Data from 9 patients with BD were collected at a different site using a Siemens Magneton PRISMA. Group comparisons include scanner as a control regressor. Scan protocols were carried out following (14), supplementary methods [3A].

### FMRI analysis – whole-brain

#### General approach

Data were pre-processed using FSL ((41), supplementary methods [3B]). Statistical analysis was performed at two levels, event-related GLM for each participant, followed by group-level mixed-effect model using FSL’s FLAME 1 (42,43) with outlier de-weighting. Whole-brain images are all cluster-corrected (p<0.05 two-tailed, FWE), voxel inclusion threshold: z < 2.3.

#### Regression designs

At the time of the decision, we looked for neural activity correlating with the utility (reward, loss) of the choice. At the time of the outcome of the gamble, we looked for neural activity related to the processing of the outcome (win/loss as continuous regressor). Decision and outcome-related activity could be dissociated due to jitter used in the experimental timing (14). As a key measure of interest, we looked at whether there was a history effect at the time of the choice (i.e., previous trial’s gamble win/loss outcome (14,44), analogous to the behavioural analyses). Full design information: supplementary methods [3C], Figure S2.

#### Group-level comparisons

We compared the low vs high MDQ groups (n=77) in whole-brain analyses. As only 22 patients with BD were available, these group comparisons were first performed in regions of interest (ROIs) derived from comparisons of the high/low MDQ groups. As exploratory analyses, BD groups were also compared at the whole-brain level.

#### ROI analyses

Mean brain activations (z-stats) were extracted for each participant. These were used to illustrate group differences and also to perform independent statistical tests (e.g., ROIs of clusters defined based on group differences of high vs. low MDQ could be used to test group differences between lithium and placebo). For this, non-hierarchical Bayesian regressions were used, also controlling for age and gender. Brain activations for the outcome history effect were correlated with the corresponding behavioural measures. For this, effects of age, gender and group (and for the patients with BD: number of days in the baseline phase pre-randomization, i.e. befor<u>e</u> the MRI scan) were first removed using regressions from both neural and behavioural measures.

## RESULTS

We recruited four groups of participants in two separate studies (Table 1). In the group of patients with BD, based on self-report scores (Altman self-report scale, quick inventory of depressive symptomatology), in the phase before the assignment to placebo or lithium, 30% scored in the mania range and 53% scored at least moderate symptoms of depression (Table 1). Similar numbers persisted throughout treatment with lithium vs. placebo (Table 1, Figure S3). Participants with BD took several medications at study inclusion (Table 2).

**Table 1.**
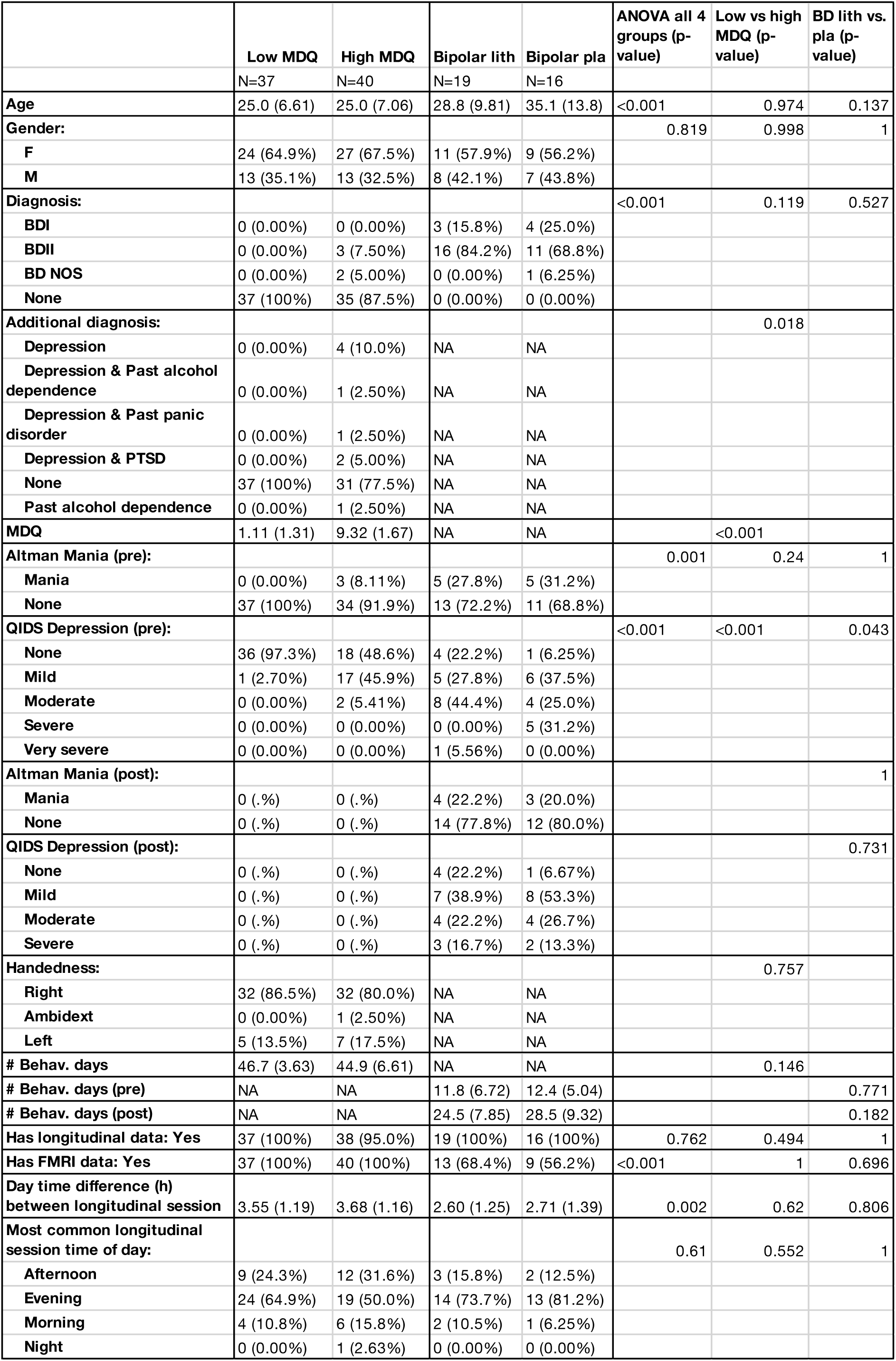
Participant demographics. Statistical tests are two-tailed p-values and refer to comparisons between the two groups of participants with low or high MDQ scores (‘Low vs. high MDQ’) and between the two groups of patients with BD randomized to lithium or placebo (‘Lith vs. pla’). Values are the mean and standard error of the mean. Abbreviations: ‘# Behav. Days’ – number of days of behavioural data available (20 trials per day), ‘# Behav. Days (pre)’ – number of days in the baseline phase for the patients with BD, ‘# PANAS days’ – number of days with mood scores (PANAS, positive affect negative affect scale, short form) available. ‘MDQ’ – Mood disorder questionnaire. ‘Has longitudinal data: Yes’ – percentage of participants from whom longitudinal data (i.e., sessions at home) were available. Diagnoses: ‘BD-I’ – bipolar I disorder; ‘BD-II’ – bipolar II disorder; ‘BDNOS’ – bipolar disorder not otherwise specified; ‘PTSD’ – post traumatic stress disorder. For the patients with BD, comorbid disorders were not measured. Note that in the low and high MDQ groups, diagnoses were only based on SCID, not on a full clinical examination. Participants completed weekly self-report scales of symptoms of mania (Altman) and depression (QIDS) at baseline (pre) and post assignment to lithium or placebo. The average scores pre (baseline) and post lithium were here categorized according to standard cut-offs (Altman: <6 for no mania, QIDS: 1-5: no depression, 6-10: mild depression, 11-15: moderate depression, 16-20: severe depression, 21-27: very severe depression). In short, lithium vs. placebo did not affect ratings of mania and depression, in line with the groups recruited here being outside major mood episodes requiring immediate treatment (see figure S3 for time course of ratings).

**Table 2.** Medication in patients with BD. At baseline, most patients were on stable doses of different medications, categorized here as: atypical antipsychotics (quetiapine, olanzapine, aripiprazole, risperidone, amisulpiride), benzodiazepine (clonazepam, lorazepam, diazepam), beta blocker (propranolol), mood stabilizer (valproate, lamotrigine), noradrenaline (NA) and dopamine (DA) reuptake inhibitor (buproprion), nonbenzodiazepine (zopiclone), sedative (promethazine), serotonin and noradrenaline reuptake inhibitor (SNRI, venlafaxine), SSRI (selective serotonin reuptake inhibitor (sertraline, citalopram, fluoxetine), tetracyclic antidepressant (mirtazapine), tricyclic antidepressant (dosulepin, lofepramine, amitriptyline), typical antipsychotic (stelazine, haloperidol).

|  | Bipolar lith | Bipolar pla |
| --- | --- | --- |
| # Participants | N=19 | N=16 |
| Medication: |  |  |
| None | 5 (8.93%) | 3 (9.68%) |
| Atypical antipsychotic | 16 (28.6%) | 6 (19.4%) |
| Benzodiazepine | 2 (3.57%) | 1 (3.23%) |
| Beta blocker | 1 (1.79%) | 0 (0.00%) |
| Mood stabilizer | 5 (9.80%) | 3 (10.7%) |
| NA and DA reuptake inhibitor | 1 (1.79%) | 0 (0.00%) |
| Nonbenzodiazepine | 1 (1.79%) | 1 (3.23%) |
| Sedative | 1 (1.79%) | 0 (0.00%) |
| SNRI | 1 (1.79%) | 0 (0.00%) |
| SSRI | 20 (35.7%) | 12 (38.7%) |
| Tetracyclic antidepressant | 2 (3.57%) | 2 (6.45%) |
| Tricyclic antidepressant | 0 (0.00%) | 3 (9.68%) |
| Typical antipsychotic | 1 (1.79%) | 0 (0.00%) |

### General performance

Participants completed longitudinal daily behavioural test sessions at home, consisting of 20 trials of a gambling task and mood self-reports. In the task (Figure 1A), participants needed to choose repeatedly between two gambles (wheels of fortune), considering the probabilities of winning or losing points and the number of points that could be won or lost. Participants in all groups performed the task well (Figure 1B), selecting options with higher values more frequently.

### Risk taking (avoidance of potential losses)

To test whether sensitivity to (i.e. avoidance of) potential losses vs. wins when gambling was reduced with a bipolar disorder gradient (low MDQ ≤ high MDQ ≤ patients with BD), we built a stochastic decision-making model that described participants’ choices as being based on the reward and loss utilities of the two options while allowing for individual differences in how people made decisions (see table S2 for model comparisons; model accuracy: 71%). The model captured participants’ sensitivity to losses (vs. wins) as a parameter (λ). We found that the higher the bipolar disorder gradient, the lower the sensitivity to losses vs wins (Figure 2Ai, Table S4A, mean =-0.27, 95%CI = [-0.49; -0.05]). This was driven mainly by a step change decrease in the group of patients with BD compared to the low/high MDQ groups, rather than a continuous linear relationship (table S4A for group comparison and continuous measure of mania symptoms across all groups). Lithium vs. placebo did not affect this (Figure 2Aii, table S4B). To illustrate the effect in a model-free way, we plotted the sensitivity of choices to the win or loss dimensions (i.e., steepness of the curve, Figure 2Aiii). This revealed that the difference between groups (group*win/loss dimension* utility bin: mean=0.33, 95% CI = [0.06; 0.61]) is driven by both an increased sensitivity to wins (group*utility bin: mean 0.24, 95% CI = [0.08; 3.99]) and a decreased sensitivity to losses (group* utility bin: mean = -0.15, 95% CI = [-0.30; -0.01]) with the bipolar disorder gradient. Alternative computational models in Table S5.

**Figure 2.**
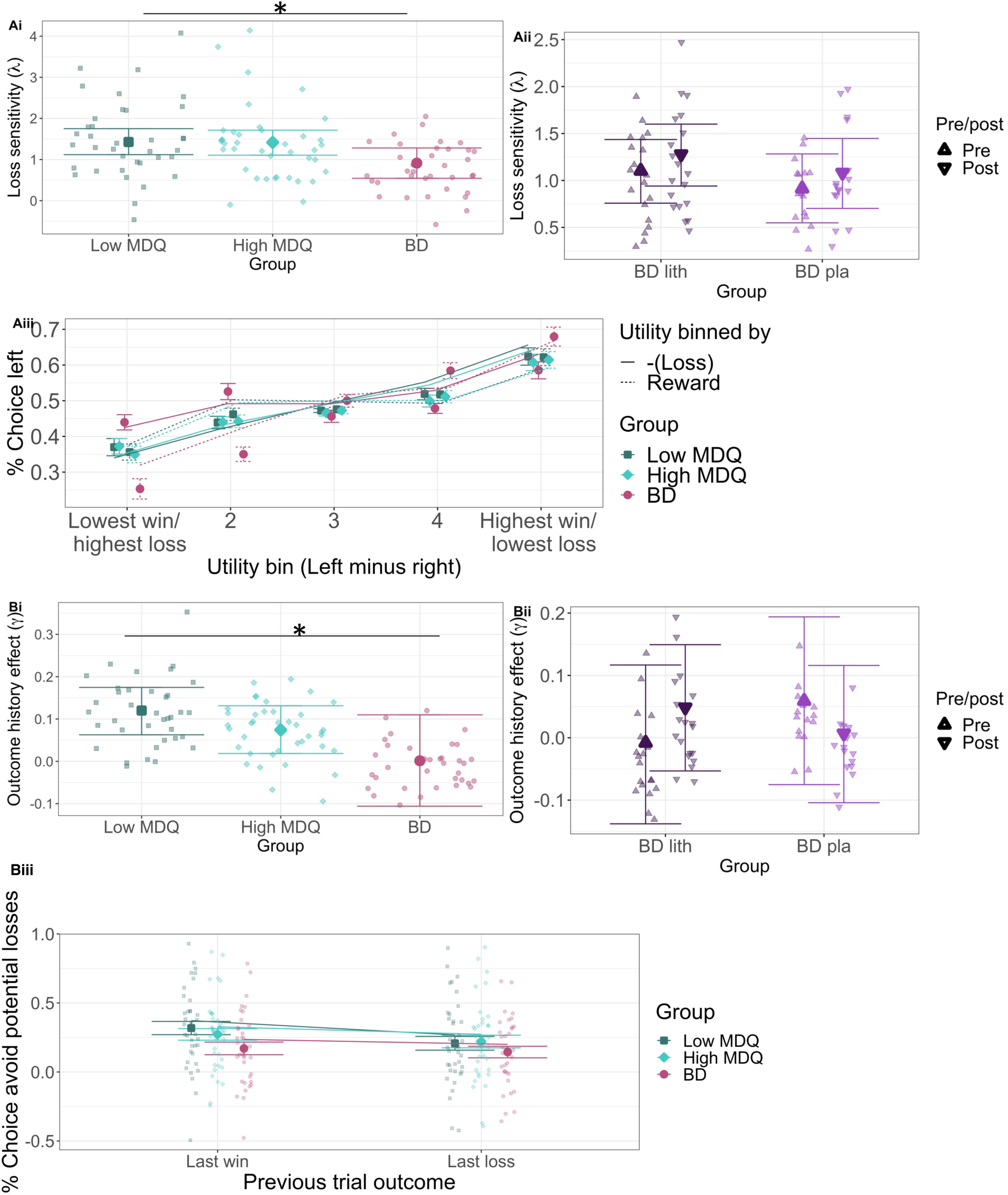
Group differences in longitudinal behaviour and mood. A) Loss sensitivity. Ai) Decreased loss sensitivity (λ, avoidance of potential losses) with bipolar disorder gradient, particularly for patients with BD. Aii) Lithium (vs. placebo) did not affect loss sensitivity (group [lithium/placebo] * time [pre, i.e. baseline/post] interaction). Aiii) Illustration of sensitivity of choices to loss/reward utility – as utility increases for the left compared to the right option, participants are more likely to choose the left option. For low/ high MDQ participants, this increase in choice probability is similar for the reward or loss dimension. In contrast, patients with BD show decreased sensitivity to losses vs. rewards (the loss curve is shallower. B) Outcome history (i.e. adaptation of risk taking to past outcomes; avoidance of potential losses after a win [rather than loss] on the previous trial). Bi) The outcome-history model parameter (γ) differed between the groups, with low MDQ participants showing the most and the patients with BD showed the least outcome history effects. Bii) Lithium (vs. placebo) did not affect outcome history effects. Biii) After a win vs. a loss on the previous trial (‘last win’/ ‘last loss’), low MDQ participants avoided losses more, while this was reduced with the MDQ gradient, so that patients with BD did not adapt their choices to past trial outcomes. A full list of comparisons of parameters for the groups is shown in Tables S4 (longitudinal data) and Table S6 (fMRI session data). Relationships between parameters measured longitudinally over weeks or in the lab during the fMRI session are shown in Table S7. ii) and iii) show conditional effects from regression models, roughly equivalent to means, controlling for regressors of no interest. Lines in Aiii and Biii show the choices predicted by the model. Participant numbers: low MDQ: 37, high MDQ:40, BD lithium: 19, BD placebo: 16.

### Outcome history effects

We next analysed how participants adapted their risk taking across trials based on win or loss outcomes in the previous trial (‘outcome history effect’). In the computational model, outcome history effects were captured as a parameter (γ) that described to what extent participants were more sensitive to (i.e. avoidant of) potential losses after a win on the previous trial. We found that the bipolar disorder gradient reduced outcome history effects (Figure 2Bi, Table S4A+S5 mean=-0.05, 95% CI=[-0.11; -0.0003], showing also a continuous effect with mania symptoms across all groups, table S4A). This was not affected by lithium (Figure 2Bii, Table S4B+S5). We can unpack this effect in the data without a model (Figure 2Biii) by focusing on the most extreme loss utility bins (if the loss utility difference is small, it will not affect choices if it is taken slightly more or less into account). If people show no outcome history effect, their choices should not change depending on the last trial’s outcome. However, the low MDQ group in fact takes the loss dimension more into account after a previous trial win vs. loss (i.e. less likely to pick options with high potential loss and more likely to pick options with low potential loss). This effect decreases with the bipolar disorder gradient (group*last loss/win: mean=0.02, 95% CI = [0.0008; 0.04]).

### Mood

Finally, an advantage of the behavioural data being collected at home was that we could relate daily mood ratings to task-based behaviour. As reported previously (3,45) and similar to other studies (2,46,47) groups differed in their instability (standard deviation) of mood: The low MDQ group showed the lowest and the patients with BD the highest mood instability (positive PANAS: mean= 0.22, 95%CI = [0.11; 0.33]; negative PANAS: mean=0.64, 95%CI = [0.45; 0.83], Table S8A, Figure S4A). Lithium did not affect instability when using our measure of standard deviation here (Table S8B, Figure S4B), though note that using a measure of Bayesian volatility, lithium has been found to increase volatility of positive mood (3). Across all groups, happiness VAS at the end of each session, compared to before was increased by overall (summed across the whole session) reward and decreased by loss outcomes (mean = 0.42, 95%CI = [0.31; 0.52]), similar to previous reports (48,49). However, this did not differ by bipolar disorder gradient (mean =-0.06, 95% CI = [-0.15, 0.03], table S8C). While mood instability differed between the groups, the impact on behaviour was distinct, with mood instability affecting the choice noisiness (the more unstable the mood, the more random the choices), without clearly affecting either loss sensitivity or outcome history effects (Figure S4C). The relationship between mood (PANAS) on the day of testing (rather than an overall measure of instability) and behaviour was not robust (table S8D). An exploratory analysis found that in the BD group, positive mood (PANAS) before the session led to reduced choice noisiness (figure S4C, stats on the regression interaction term BD gradient x PANAS predicting choice noisiness: mean: 0.18, 95%CI: [0.01; 0.35]).

### Neural results

Neural data were available for 77 volunteers and 22 patients. Across volunteers, brain activations to reward and loss utility during decisions (Figure 3A) and at the receipt of outcomes (Figure 3C) activated brain evaluation networks, including ventromedial prefrontal cortex (vmPFC), ventral striatum, dorsal anterior cingulate cortex (dACC), insula (Table S9). Next, we tested whether, related to the outcome history effect, there was brain activity when participants made a choice that correlated with the previous trial’s outcome. Indeed, we found that activity in a network including the ventral striatum, vmPFC and medial frontal pole (FPm) related to the outcome of the previous trial, i.e. increased activity the more positive (and less negative) the previous trial’s outcome (Figure 3B, Table S9).

**Figure 3.**
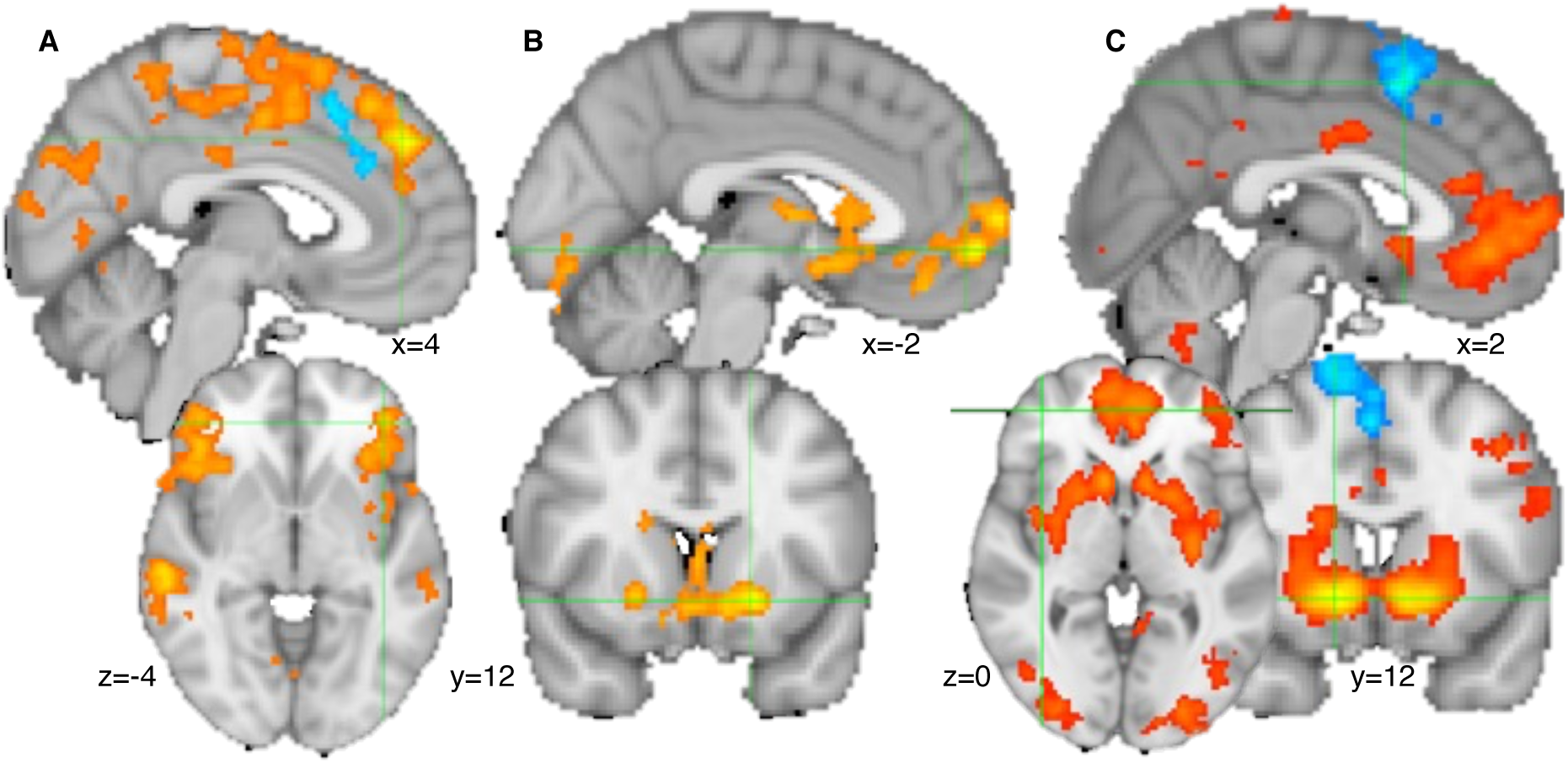
Neural activity during gambling. A) At the time of the decision a wide network of areas activated with relative (chosen minus unchosen) reward utility (orange), while loss relative utility activated the anterior cingulate cortex (blue). B) At the time of the decision, the last trial’s outcome (points won or lost) activated areas including vmPFC and ventral striatum (orange). C) At the time of the outcome (win or loss received), the outcome (points won or lost) activated areas including vmPFC, FPm, and ventral striatum (red/orange) and deactivated the pre-supplementary area. All results are cluster-corrected at p<0.05, two-tailed, with inclusion cut-off z>2.3. See Table S9 for the full list of results. Data were combined across both volunteer groups (low and high MDQ). Participant numbers: low MDQ: 37, high MDQ: 40.

Next, we compared the low and high MDQ groups. Activity for the previous trial’s outcome was higher for the low MDQ vs high MDQ group in FPm (Figure 4Ai-ii, Table S10, p=0.038, whole-brain cluster corrected). In other words, while all participants showed activity in vmPFC/FPm, in low MDQ participants the cluster extended further into FPm. Moreover, there was a correlation between the neural signal for the previous trial’s outcome and the behavioural outcome history effect: the stronger the activity for the last trial’s outcome in this area, the stronger the behavioural outcome history effect (Figure 4Aiii, r=0.24, p=0.017, partial correlation after correction for control variables and group; without correction: r=0.28, p=0.005; test performed as robust regression, controlling for outliers: 95% Bayesian CI = [0.03; 1.52]). Lithium vs. placebo participants’ activity did not differ in this area (mean=0.64, 95% CI = [-0.23; 1.44]).

**Figure 4.**
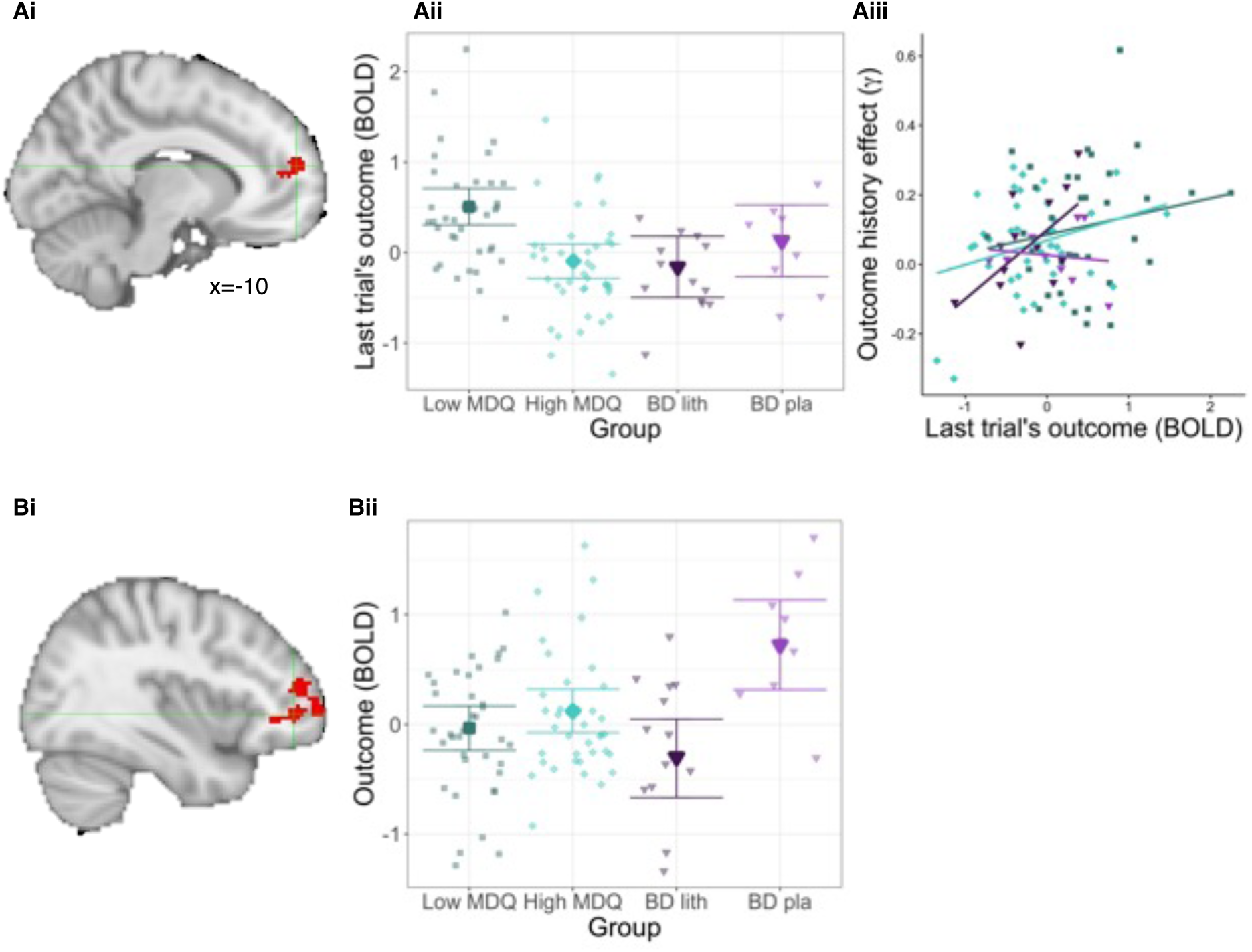
Group differences in brain signals. A) Differences between the low and high MDQ groups for the outcome history effects. Ai) Activation with last trial’s outcome at the time of the current trial’s decision differed between the low and high MDQ groups in the medial frontal pole (FPm; x=-10, y=56, z=16; p=0.038, n=77, cluster-corrected, Table S10A. In the low MDQ group, the activation with the last trial’s outcome that is found across both groups (Figure 3B) extends further dorsally. Aii) This group difference was driven by the low MDQ group showing stronger activation than the high MDQ group in FPm (Figure shows conditional effects from regression model, roughly equivalent to means, controlling for regressors of no interest). There was no significant difference between activations comparing lithium and placebo groups (-0.30, 95%CI: [-0.73; 0.17]). Aiii) This FPm activity correlated with the longitudinally measured outcome history parameter. Related whole-brain results shown in Figure S5. Colours match those of groups in B. B) Exploratory whole-brain group differences in the patients with BD for gamble outcome signal (lithium vs. placebo). Bi) Outcome related activity differed between the placebo and the lithium participants in an area including dorsolateral prefrontal cortex and lateral frontal pole (whole-brain cluster-corrected, Table S10B). This effect is illustrated in Bii). Participant numbers: low MDQ: 37, high MDQ:40, BD lithium: 13, BD placebo: 9.

As exploratory analyses (due to low sample sizes in BD groups for the MRI scan), we next compared lithium vs. placebo treatment at the whole-brain level. We found that patients receiving placebo had stronger activity related to the outcome of gambles in an area spanning dorsolateral prefrontal cortex (dlPFC, area 46) and lateral frontal pole (Figure 4B, Table S10B, p=0.009). We also tested whether the gamble outcome activation related to the behavioural outcome history effect, finding that interestingly it did (Figure S5), in an mFP area overlapping with the area of group differences identified above, though not in the dlPFC area of group differences between lithium and placebo (Figure 4A).

## DISCUSSION

We designed a study to test the computational and neural correlates of adaptations of risk-taking to gains and losses in bipolar disorder (BD), in risk of bipolar disorder and treatment with lithium. We included participants along a gradient of bipolar disorder ranging from volunteers with low risk of BD (low MDQ group), to volunteers with high risk of BD, to patients with diagnosed BD. In the patients, we tested the effect of lithium treatment in a placebo-controlled double-blind design. We measured how much participants adapted their risk-taking following reward outcomes in a risky decision-making task (‘outcome history effects’). We measured behaviour both longitudinally over up to 50 days and during a brain imaging (FMRI) session. We found that the low MDQ group showed an ‘outcome history effect’. Specifically, after a win on a trial, they were more risk averse (avoiding potential losses). This was reduced across the bipolar disorder gradient (lowest risk aversion adaptation in patients with BD). Neurally, outcome history was related to the representation of past information in a large network including ventromedial prefrontal cortex (vmPFC) and medial frontal pole (FPm). In low MDQ volunteers, this brain signal extended further dorsally into FPm compared to the high MDQ scorers and this was correlated with risk adaption behaviour.

Decreased loss sensitivity and reward hypersensitivity have been suggested as central to BD (39,40) (50,51) and may drive risky or impulsive decision making. Our findings of decreased sensitivity to potential losses (vs wins) with BD gradient are in agreement with this. This effect showed a step change between the volunteer and the patient groups, rather than a continuous effect across the gradient. Then, we went further looking at adaptation of risk taking to past outcomes. We found that volunteers with presumed low risk of bipolar disorder (low MDQ) showed sequential dependencies between their choices and previous trials’ outcomes, avoiding potential losses after a win on the previous trial (‘outcome history effect’), as similarly recently reported in a go/no go decision-making task (52) . This was not strictly rational in our task since outcomes for gambles across trials were independent (10). However, this kind of behaviour observed in the lab may be functionally appropriate in more naturalistic environments (38,53–55) and thus reflect prior beliefs participants have about reward distributions (e.g. non-independence between trials). For example, in natural environments, which are experienced continually rather than in discrete trials and in which different types of rewards (e.g. food, water) need to be accumulated or a homeostatic setpoint needs to be reached, it would make sense to adapt behaviour according to previous outcomes (56–60). The influence of past losses (vs wins) was lower in the high vs low MDQ group and lowest in patients with BD (i.e. the pattern showed a continuous gradient, also captured as a linear relationship to mania scores across all groups, rather than a step change from volunteers to patients). Reduced homeostatic behaviour of this kind could lead to unstable moods since in the healthy population mood has been found to be regulated through behaviour (8). Relatedly, in patients with BD, purposefully regulating behaviour during the prodromal periods has been shown to reduce the risk of relapse (61). However, in our study, links between ratings of mood and behaviour were weak and so this suggestion remains speculative. Future studies could measure mood over longer timescales, more frequently than done here and using a more naturalistic task. We also note that our findings diverge from previous findings (4) of a stronger impact of previous rewards (and associated emotions) on the perception of outcomes in a study including a participant sample not specifically selected for BD diagnosis or risk of bipolar disorder, but completing the Hypomanic Personality scale (62) after the task.

We focused on whole-brain analyses for the low/high MDQ volunteer sample due to the larger sample size compared to the patient study. Decision-making and the processing of outcomes produced a typical pattern of activation (63–66) in areas including dorsal anterior cingulate cortex, striatum and vmPFC. However, there were no group differences in any of these signals, matching our behavioural results of an absence of differences in general ability to make decisions or sensitivity to rewards vs. losses per se in the low vs. high MDQ groups. We next looked for brain activity related to the modulation of risk taking with ‘outcome history’. We found that at the time when people made decisions, there was activity representing the last trial’s outcome in an area spanning vmPFC to FPm. This is similar to previous findings in a learning context of between-trial activities (14,44,67). This gamble outcome activation was related to the behavioural outcome history effect across participants. This signal extended more dorsally into FPm in low MDQ volunteers. Furthermore, the stronger this signal, the stronger the modulation of risk taking by outcome history. As such the influence of outcomes on decision making may be a feature of risk for bipolar disorder which involves the FPm. This adds to previous work linking BD to changes in reward related signals in ventral striatum and OFC (19,68) and changes in connectivity between striatum and PFC (17,18). In this region, lithium did not affect brain activity, suggesting that its mechanism of action may not involve direct modulation of vmPFC value weighting.

In an exploratory analysis, we compared the brain activity of patients with BD randomised to lithium or placebo. Patients given placebo showed larger outcome-related activity in dorsolateral prefrontal cortex. Yet at the same time, lithium did not change behaviour. dlPFC signalling has largely been associated with regulation of mood and reward-related behaviour. Previous work in bipolar disorder has showed altered patterns of both vmPFC and dlPFC activity. In particular, Mason et al. (17) reported that while controls activated dlPFC more to rewards of high probability, patients with bipolar disorder showed greater dlPFC to low probability (more risky) rewards. As such, our preliminary findings suggest that lithium may modulate a key component of frontostriatal circuitry important for effective decision making. Previous work in healthy volunteers also reported an effect of lithium on reward related signals in the ventral striatum which wasn’t detected in the current study(69).

The current work has a number of limitations. Our sample size was low for the comparison between lithium and placebo fMRI responses, which may have affected our statistical power for key comparisons. It is also relevant that we saw no effect of lithium on the clinical questionnaires included in this study. However, this is consistent with the characteristics of the sample recruited here, where current symptoms were largely residual (i.e. outside of an acute episode). Furthermore, lithium is largely used for relapse prevention rather than acute treatment of mania or depression (70) which could not be tested in the short timescale of the current investigation. Data across a large number of tasks and measures were also completed as part of these studies, and analysis is still ongoing. These complete results may shed light on the overall effects of bipolar disorder risk and treatment on different facets of mood and cognition. While we pre-registered our lithium trial (2014-002699-98), we did not pre-register our specific hypotheses for this part of the analysis. While we found an expected value signal (chosen minus unchosen value) in a typical ‘negative value’ network including the dACC, we did not find a ’positive value’ signal in a typically expected area like the vmPFC. This is unlikely to be due to signal drop out as vmPFC showed activation with reward outcome and an outcome history signal at the time of choice. This result is reminiscent of our previous findings (14), where it was interpreted as possibly due to the integration of an aversive dimensions (there: effort) with reward, rather than only integrating two positive dimensions (e.g. reward probability and reward magnitude). Similarly, here, participants were faced with a negative dimension, i.e. monetary loss.

Our results highlight the importance of considering rewarded decision-making and related neural activity to understand symptoms of bipolar disorder and the stabilising effects of lithium.

## Contributions

JS: Conceptualization, software, formal analysis, writing, original draft, reviewing and editing

PP: Investigation, writing, reviewing and editing

NN: Conceptualization, investigation, reviewing

NK: Conceptualization, reviewing and editing

LZA: Investigation, writing, reviewing and editing.

MFSR: Conceptualization, design, reviewing and editing.

ACN: Funding acquisition, conceptualization, supervision, design, reviewing and editing.

PJH: Funding acquisition, Conceptualisation, recruitment, reviewing and editing

JG: Funding acquisition, conceptualisation, design, supervision, reviewing and editing

KS: Investigation, project administration, reviewing and editing

CJH: Funding acquisition, conceptualization, design, supervision, reviewing and editing

## Code and data availability

Code and anonymized (behaviour, selected demographics, brain activity from regions of interest) or group level (whole-brain FMRI) data will be available on osf.io upon publication. Access for reviewers (link to be replaced with public one upon publication): https://osf.io/ychbf/?view_only=0a6fce5cc7e74e9c88c542b6aa7d8680

## Data Availability

All data produced in the present study are available upon reasonable request to the authors.

## Supplementary methods

### [1] Additional participant information

#### [1A] Exclusion criteria

##### Healthy volunteers with low and high mood instability

Common inclusion criteria across both groups were: over 18 years old, no current medication (other than contraceptive pill), no use of antidepressants, antipsychotics, lithium or anticonvulsant medication in the last 6 weeks, no contraindication to MRI or MEG. Participants were recruited based on their scores of mood instability (MDQ). The questionnaire includes first questions describing symptoms of bipolar and second a question about whether the different symptoms occurred at the same time. Participants were included in the low MDQ group if their number of reported symptoms was 5 or less. Participants were included in the high MDQ group if they reported 7 or more symptoms and report that they have happened at the same time. Additionally, in the low mood instability group, we excluded participants with a current or past diagnosis of an axis 1 psychiatric disorder (assessed using DSM-IV interview) or a first degree relative with bipolar disorder. In the high mood instability group, we excluded participants with a current or past diagnosis of an axis 1 psychiatric disorder other than bipolar disorder I or II, major depression or anxiety disorders.

##### Participants diagnosed with bipolar disorder

Inclusion criteria: over 18 years old, meeting criteria for BDI, BDII or BDNOS as assessed using the Structured Clinical Interview for DSM-V Axis I Disorders (SCID-I), clinically significant mood instability (established through interview), not currently suicidal (currently suicidal assessed as a score of ≥ 4 on the C-SSRS (Columbia Suicide Severity Rating Scale Score (8)), no counterindications to lithium (assessed through pre-treatment tests including renal, cardiac, thyroid and parathyroid functions), not currently taking any psychotropic drugs that could not be withdrawn, not requiring acute treatment so that placebo would be inappropriate, participation in a previous research trial in the past 12 weeks. Participants with counterindications to MRI scanner were included in the behavioural part of the study.

##### All participants

When inspecting the PANAS daily mood ratings, we noticed that for some participants on some days, every single response (to all positive and negative items was zero). This suggests a technical problem. We set the data from these days to zero. There were 25 participants (across all groups) for who this happened for more than 10% of measurements. Because of using hierarchical models, we could include most participants for all analyses. Specifically, we could include all participants that did not have a standard deviation of zero for the mood measures (this excluded 1 participant for the analyses for negative PANAS, 0 for positive PANAS and 2 for the analyses of changes in mood before and after the task).

#### [1B] Larger study – full information

##### Volunteers

The data presented here was part of larger study (CONBRIO, Collaborative Oxford Network for Bipolar Research to Improve Outcomes: The cognitive neuroscience of mood instability; Cognition and Mood Evolution across Time (COMET) – MSD-IDREC-C2-2014-023). Participants who expressed interested in the study were given an electronic version of the information sheet and the Mood Disorder Questionnaire (MDQ). If they scored in either the ‘low MDQ’ or the ‘high MDQ’ category they were invited for a first study visit.

During the first study visit, inclusion and exclusion criteria were checked. Participants were also screened for psychiatric disorders using the Structured Clinical Interview for DSM Disorders (SCID) (9). Participants completed several questionnaires, including Barratt Impulsiveness Scale (10), Sleep Condition Indicator (11), Maclean Screening Instrument (12), Affective Lability Scale (13), Affect Intensity Measure (14). Participants were set up with and instructed how to use devices to measure their activity (GeneActiv watch and FitBit) over the ten weeks study period. They were given an iPad mini and trained on four cognitive tasks: ‘wheel of fortune’ – risky decision making (presented here), ‘guess the gap’ – performance learning, ‘fractals’ – stimulus-outcome learning, ‘whack-a-t’ – implicit spatial learning.

Over a period of ten weeks, they were asked to complete these tasks five times a week and to wear the GeneActiv and FitBit devices as much as possible. They were also asked to complete clinical questionnaires (Quick Inventory of Depressive Symptomatology (15), Altman self-rating mania scale (16), Generalized Anxiety Disorder-7 (17), EuroQol-5 [health-related quality of life] (18)) on the True Colours mood monitoring system (19) once a week. They were also asked to stay within the recommended daily alcohol intake levels throughout.

In the beginning of the ten-week period (weeks one or two) and in the end (weeks nine and ten), participants attended an MRI scan and a MEG scan. Due to MEG scanner downtime, only 24 participants received the MEG scan. During both MRI scans, resting state and structural data was obtained. At the first scan, additional the ‘wheel of fortune’ was measured. At the second MRI scan, ‘fractals’ and ‘guess the gap’ was measured, as well as diffusion tension imaging and fluid-attenuated inversion recovery.

##### Participants diagnosed with bipolar disorder

Participants first took part in a screening visit in which inclusion criteria were checked and informed consent was taken. Using the SCID-I, a diagnosis check was done. In addition, demographic and clinical information was obtained, including duration of illness, previous use of psychotropic medicines, family history of mood disorders, presence of comorbid borderline personality disorder and attention deficit hyperactivity disorder and current suicidal ideation, concomitant medication and substance use and a physical examination. If blood samples have not been taken as part of routine monitoring, they were taken at this visit. Two sets of samples were taken, one was sent to the pathology lab for analysis and the other was retained as replacement for samples lost/damaged in transit and for storage for future research. Tests included urea and electrolytes, full blood count, fasting blood glucose, glycosylated haemoglobin (HbA1c), blood lipid profile, LFTs, T4, T3, TSH, thyroid antibodies, PTH, vitamin D, eGFR, Cystatin C and NGAL and inflammatory markers CRP and IL-6. A sample was taken to measure calcium level using the InSight™ Electrolyte Analyser located in the NIHR-CRF. Weight/BMI, pulse and blood pressure were also recorded, and an ECG was performed. Participants were given an iPad mini and trained on the same cognitive tasks as the healthy volunteers described above. They were also set up with the True Colours system to rate weekly mood and on Mood Zoom (20) to rate daily mood. Participants were also given activity monitors. They were also given saliva swabs.

Before being randomised to lithium or placebo, all participants completed two weeks of daily cognitive tasks, mood and activity measurements at home (though some participants completed up to 30 days due to logistic challenges). Then, they were randomised and performed six weeks of cognitive tasks, mood and activity measurements.

In the beginning of the six weeks period (week one or two), they completed an MRI and a MEG scan using the same scans as described for the healthy volunteers.

Randomisation: The first 10 participants were fully randomly assigned to avoid predictability, while for subsequent participants, an algorithm was used to minimize differences in age (<25 or >25 years) and gender between the two groups. In the lithium group, participants were titrated to doses producing plasma levels of 0.6-1 mmol/L (see supplementary methods 1C for dosing details).

#### [1C] Lithium dosing information

In the lithium group, participants were prescribed an initial dose of 400g/day, unless there was a clinical indication to start at a lower dose. During this phase, participants attended brief assessments at 4-days, 8- days and between 2 and 3 weeks post-randomisation to review lithium levels by a psychiatrist (if lithium level ≤ 0.3 mmol/L, dose increased to 800mg/day; if lithium level between 0.4 to 0.5 mmol/L, dose increased to 600mg/day; if lithium level 0.6 -1.0mmol/L, continued current dose; if lithium level ≥ 1.0mmol/L, decrease dose by 200mg/day or 400mg/day as found appropriate by psychiatrist), receive additional supplies of lithium/ placebo as needed and were asked about adverse events. Participants took part in one neuroimaging session in week 3 or 4. During the trial, participants were asked to complete the cognitive tasks daily.

### [2] Computational modelling, additional information

#### [2A] Decision making model validation

We validated our computational models using simulations (21,22). We simulated 400 participants with parameter values (mean and standard deviations) drawn from a uniform distribution in the 95% range of the parameters for real individual participants. For each participant, we simulated 47-50 sessions (uniform distribution). Parameters for single sessions were drawn from normal distributions of simulated participants’ means, standard deviations and linear effects of days. Simulated data was then fitted using same approaches as above. To speed up fitting of data, variational Bayesian approximation (23) was used unless control indices (pareto smoothed importance sampling, khat >0.7 (24)) suggested unsuccessful fitting even after increasing number of samples and decreasing tolerance, in which case sampling was used. When fitting the models, initially, 4 chains, with each 15,000 iterations were drawn and the target acceptance rate (adapt_delta, (25)) was set to 0.85. Whether models had been fit appropriately was checked using a criterion of R-hat (measure of mixing of chains) < 1.1 and absence of divergent samples. If these were not fulfilled, number of iterations were increased by 50% and adapt_delta was increased towards 1 (by 50% of distance from 1). This was repeated until all models converged.

To validate the model, we then checked the correlations between true and fitted values for mean and standard deviation of parameters across individual subjects (table S1).

#### [2B] Alternative decision-making models and model comparison

In addition to the models in the main text (section Computational models – decision making), we also considered two other classes of models (see table S2 for full list). First models that incorporated probability and magnitude distortions according to prospect theory (26) (class ‘M2’):

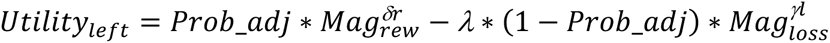

Where

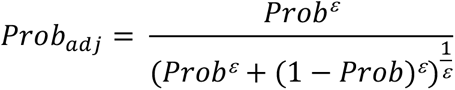

Here, ε is the probability distortion; δr is the distortion of the reward magnitudes; δl is the distortion of the loss magnitudes and λ is the weighting of the loss (scaled mag * scaled prob).

We also fitted further versions of this model, leaving out the probability distortion, the loss scale or the magnitude distortions.

Of note, due to there only being 20 trials available per session, not all of these models could be fitted. Specifically models that contained probability distortion could not be fitted and models with a loss weight in addition to exponential scaling of loss could not be fitted.

In these models, the outcome history effect was initially included in the exponential of the loss magnitude:

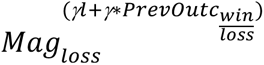

However, we noted that, potentially due to the difficulty of estimating parameters that are used as an exponent, parameter recovery for the outcome history effect in this model was not very good. Therefore, we also included the outcome history parameter as a linear weight of the exponentially distorted magnitudes.

The second set of models allowed participants to differ in their relative weighting of expected value, variance and skew (27) (class ‘M3’):

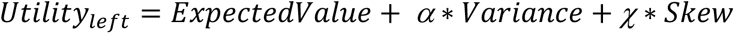

Where:

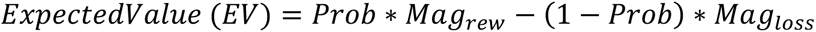

(given that Mag_loss_ is a positive number).

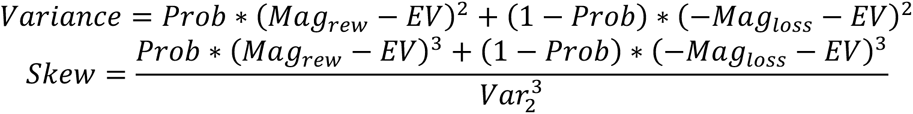

Again, we fitted this model also without the weighing for skew or without the weighing for variance. In this model, outcome history was captured as impacting the weighing of variance or skew or both.

To compare models, we used the Akaike Information Criterion (AIC) (28), which combines the log likelihood with the number of model parameters to avoid selection of over-parameterized models. Models were fit across all sessions from each participant, including for each parameter (other than outcome history effects, see table S5) a linear effect of day:

Parameter_t_ = Parameter_t0_ + day_effect*current_day

AIC values were summed across participants. Participants for whom not all models could be fit were omitted (n=2).

#### [2C] Bayesian models – additional information for standard settings

Regression models were computed with the BRMS toolbox (29) which uses the Bayesian programming language Stan (30). The key advantages of the Bayesian approach are: priors can be defined to ease fitting, particularly when little data is available (as here only 20 trials per session); models can be hierarchical and account for individual differences in each parameter (i.e. taking into account data consistent of within and between subject measurements, e.g. several data points per person and several subjects); variability in measurements across people can be taken into account.

##### Linear non-hierarchical regression

All regression estimates (parameters) were given flat priors for all parameters and 5000 iterations for each of four chains were drawn (target acceptance rate, adapt_delta = 0.9). Model fit was checked using criterion of Rhat <1.1 and the absence of divergent samples (25). If models did not converge, iterations and adapt_delta were increased step-wise, up to a max of 25312 iterations and adapt_delta = 0.991. If fitting was then still not successful (only the case for Prospect Theory models, class M2, listed in [2B] above), the sessions were left out from model comparisons.

##### Linear hierarchical regressions

To ease fitting (31), all regression estimates were given weakly informative priors, normal(0,5). 6,000 iterations for each of 4 chains were drawn (target acceptance rate, adapt_delta = 0.9). Model fit was checked as above for non-hierarchical models. Significance follows the standard definition of the Bayesian 95% Credible Interval not including zero. To compare individual groups, the same model was fitted with group as an unordered factor and posthoc tests were then done using the emmeans package (32) (again using 95% Credible Intervals to define significance). Results of regressions are illustrated as conditional effects, i.e. all other variables are set to their mean. We computed mean parameters for individual participants to relate to neural activity.

##### Computational decision-making models – prior settings

For each parameter weakly informative priors were specified for models for the longitudinal or the FMRI data for each session (longitudinal) or person (FMRI): inverse temperature (β): cauchy(5,3), weighting of the loss utility (λ): cauchy(-1,1), the impact of the previous trial’s win/loss on the weighting of the loss utility (γ); group level standard deviations: inverse temperature (β): cauchy(0,3), all other parameters: cauchy(0, 1).

When fitting the decision-making models, number of iterations and adapt_delta were increased until fit indices suggested appropriate fit, as described above.

##### FMRI

Computational models were fitted as for the longitudinal data, i.e. first separately for each individual participant before then comparing the computational model parameters across groups using non- hierarchical models (as one FMRI session per person).

#### [2D] Regressions relating mood, task outcomes and behaviour

We used hierarchical regression models to test for group differences in the impact of task outcomes on mood:

Mean: Happiness (post minus pre) ∼ 1 + Task outcomes* group + Task outcomes + group + day + Age + Gender + (1 + day + outcome | ID)

And error term: sigma ∼ 1 + group + age + gender + (1|ID)

Where outcome was either the total wins in the daily task, the total losses or the total wins minus losses. Group was coded as monotonic factor.

In addition to the happiness VAS that was measured before and after the task, we also measured mood using a more detailed questionnaire (PANAS-SF) before the task. We used this to replicate previous findings (1,20,33,34) of mood instability related to bipolar disorder (with hierarchical models):

Mean: PANAS ∼ 1 + day + group + age + gender + (1 + day |ID)

Error term: Sigma ∼ 1 + group + age + gender + (1|ID)

Where PANAS was either the positive or the negative PANAS score.

### [3] MRI scan

#### [3A] MRI acquisition sequences

Scan protocols were similar across both sites and differences are highlighted. T1-weighted structural images were acquired with the settings TR=3 sec, TE=4.71 msec (4.65ms for second site [some bipolar patients]), TI (inversion time) = 1.1 sec, 1x1x1 mm voxel size, 256x176x224 mm grid, flip angle = 8°, phase-encoding direction = R-L, GRAPPA (Generalized autocalibrating partially parallel acquisition) = 2. Functional images were acquired using a Deichmann echo-planar imaging (EPI) sequence with TR=3 s, TE=30 ms, 3x3x3 mm voxel size, 87° flip angle, 30° slice angle and z-shimming to reduce signal distortions as well as dropout in medial orbitofrontal areas (35). A fieldmap with dual echo-time images (TE1 = 5.19 ms, TE2 = 7.65 ms, whole brain coverage, voxel size 3.5 × 3.5 × 3.5 mm) was obtained for each subject to allow for corrections in geometric distortions induced in the functional images.

#### [3B] FMRI preprocessing

We used FSL (36) version 6.00 for standard image preprocessing and analysis (suppl. Methods [4B]. We used FSL’s BET (37) on the high-resolution structural MRI images and fieldmaps images to separate brain matter from nonbrain matter. We used the structural images to register functional images in MNI space using nonlinear registration as implemented in FNIRT (38). Functional images were corrected for motion using FSL’s MCFLIRT (39), corrected for geometric distortions using FSL’s FUGUE (FMRIB’s Utility for Geometrically Unwarping EPIs) and spatially smoothed with a Gaussian kernel of 5mm full-width half-maximum. Finally, images were then high-pass filtered with a 3 dB cutoff of 100s.

#### [3C] FMRI analysis

Data were pre-whitened before analysis (40). The fMRI design was as follows (see Figure S2 for design correlation matrix): We included four boxcar regressors capturing the different phases of each trial: the decision phase (aligned to the onset of the decision phase and lasting until participants could make a choice), the spinning phase (aligned to when the indicator on the chosen wheel of fortune started moving and lasting until it stopped), the outcome phase (the time the outcome was shown to participants and lasting until it disappeared from the screen) and the total score phase (aligned to when the screen with the total score was shown and lasting until it disappeared). Here the decision and the outcome phase are the main phases of interest, the others are included as control regressors. We also included parametric boxcar regressors aligned to same onsets as the phases described above, but with duration one second. All regressors were z- score normalized within each participant. In the decision phase, we included separate regressors for the reward and loss utilities (i.e. probability x magnitude) of the chosen minus the unchosen options, a regressor for last trial’s outcome (win loss, including the magnitude therefore, e.g. +10 or -20), as well as participants’ log-transformed reaction time as a control regressor. In the outcome phase, we included a regressor indicating the current trial’s outcome (win/loss, including the magnitude thereof). As control regressor we also included the total score phase with total scores as parametric value. All regressors were convolved with a double-gamma hemodynamic response function.

### [4] Bayesian mood instability models

Pulcu et al. (1) proposed a model of mood variations that captures simultaneously variability in mood ratings and drifts (volatility) in the mean mood ratings. We adapted this model here (simplifying due to less data being available than in Pulcu et al., that neither volatility nor standard deviations changed over time and fit to both positive and negative PANAS simultaneously). The same model also captured relationships between PANAS standard deviation and behaviour. The key equations of the model included:

PANAS_ratings[t]_ ∼ normal(PANAS_t_,PANAS_sd_)

PANAS_t_ ∼ normal(PANAS_t-1_, PANAS_volatility_)

Where volatility and standard deviation of PANAS ratings were shared across positive and negative PANAS. In contrast, PANAS values (PANAS_t_ above) were captured separately for positive and negative PANAS. Behaviour ∼ normal(b_0_ + b_PANAS_sd_*PANAS_sd_ + b_testing day_*testing_day, behaviour_sd_)

The model was fit to data from all participants who had at least 5 data points available for all measurements and standard deviations of both positive and negative PANAS above 0 (i.e. who did not always report exactly the same mood). Models were fitted as hierarchical models (mixed effects models), with group level parameters (mean and standard deviations) fitted for PANAS_sd_, PANAS_volatility_, b_testing_day_ and behaviour_sd_. Parameters for individual participants were then drawn from the thus defined normal distributions. PANAS_t_ for positive and negative PANAS was fitted as one parameter per person per day (with the temporal order constraints as described in the regressions above). All parameters were given priors normal(0,1), for constraint parameters (i.e. standard deviations and volatility), log transformations were used.

To test how predictive mood instability was of group membership, we first fitted a simpler model of only the PANAS scores, excluding the behaviour, separately for each person. We then used the PANAS_sd_ and PANAS_volatility_ to predict group membership in a leave-one-out cross-validation procedure, each time fitting a model of the form:

Group ∼ 1+ PANAS_sd_ + PANAS_volatility_

The model was fit to training data (i.e. all participants apart from one) and used to predict the test data (the left out participant). We trained models separately predicting low vs. high MDQ and predicting all three groups.

---

**Figure S1.**
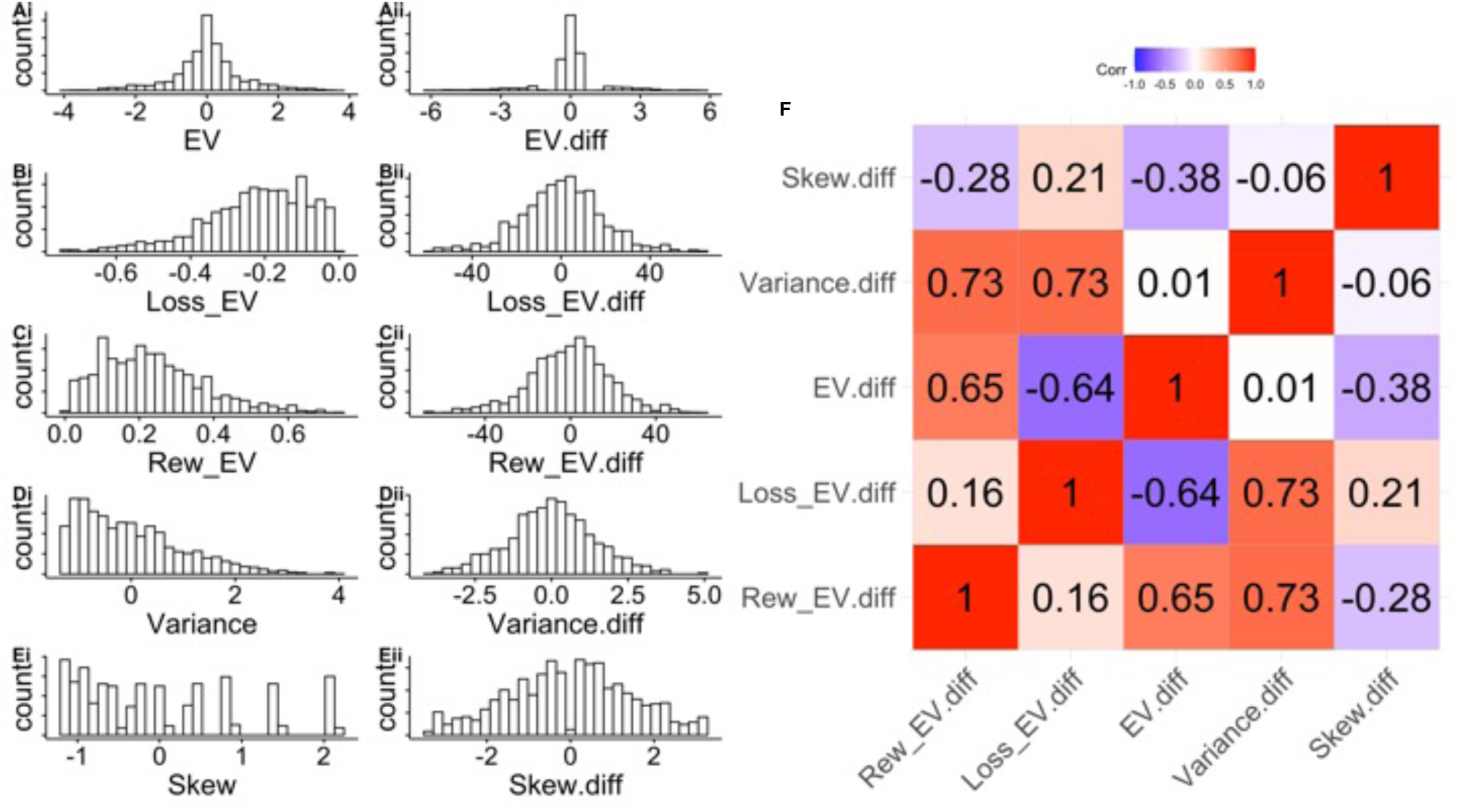
Illustration of task schedule. (related to Figure 1). A)-E) Distributions for total expected value or ‘utility’ (EV, i.e. for each option: probability*reward magnitude + (1-probability)*loss magnitude, assuming loss magnitude is coded as negative number), loss expected value ((1-probability)*loss magnitude), reward expected value (probability*reward magnitude), variance and skew across all trials of the experiment (see supplementary methods [2B]). i) shows the distribution of these values across all options and ii) the distribution of left minus right (‘diff’) options. F) Correlation between the task properties. Of note, as expected, variance and total expected value are highly correlated with both reward and loss expected value differences.

**Figure S2.**
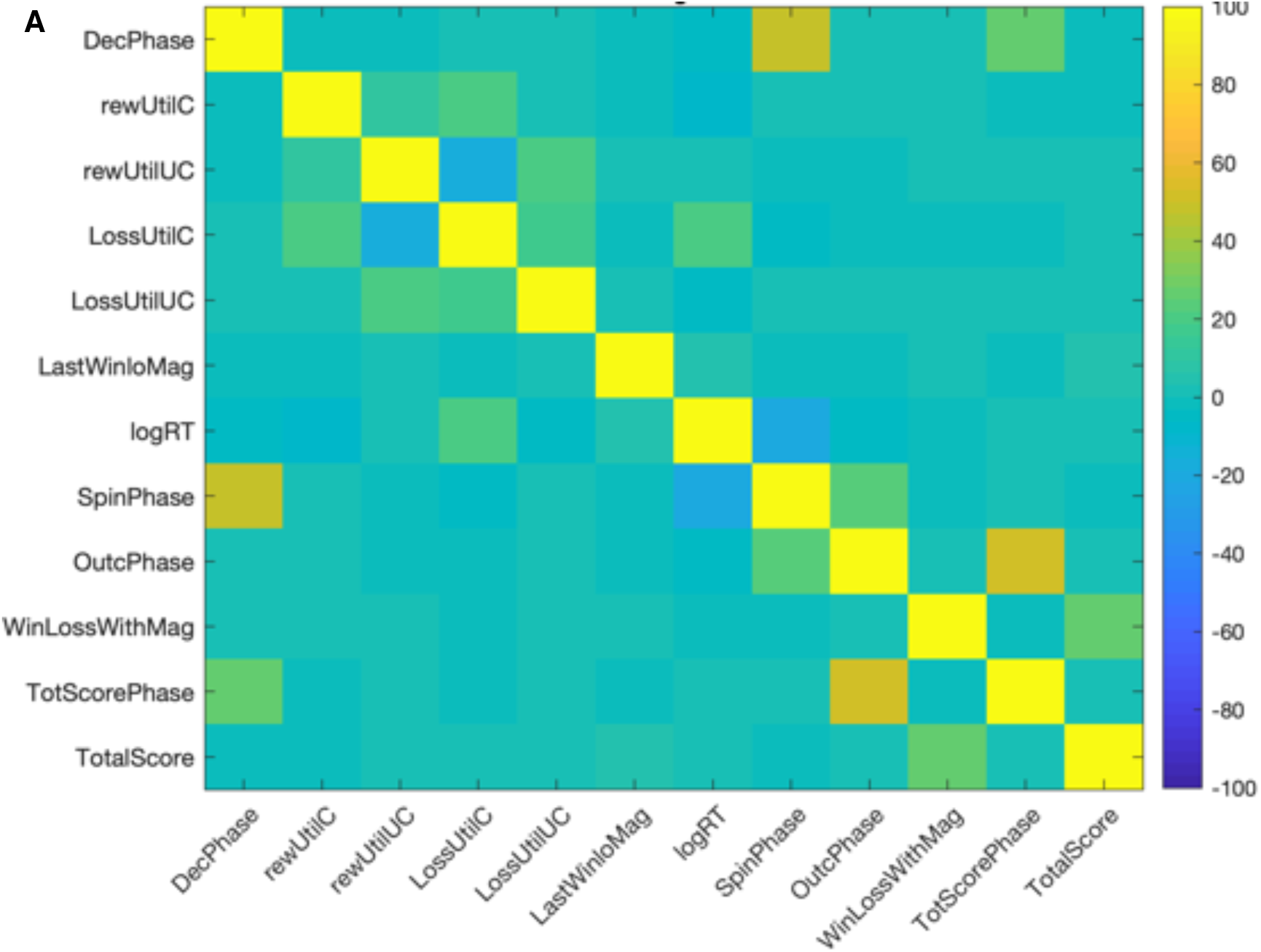
FMRI designs. Correlations of the haemodynamically convolved regressors for FMRI design 1 (A) and design 2 (B). No value regressors exceeded correlations of r>0.5 with any other regressors or confounds. Abbreviations: chosen reward utility (rewUtilC), unchosen reward utility (rewUtilUC), chosen loss utility (lossUtilC), last trial’s outcome, i.e. points won or lost, e.g. +10 or -20 (LastWinLoMag), current trial’s outcome (WinLossWithMag), relative reward utility (rewUtilCmUC), interaction between last trial’s outcome and the current trial’s loss utility (LastWinLoMagxLossUtilCmUC).

**Figure S3.**
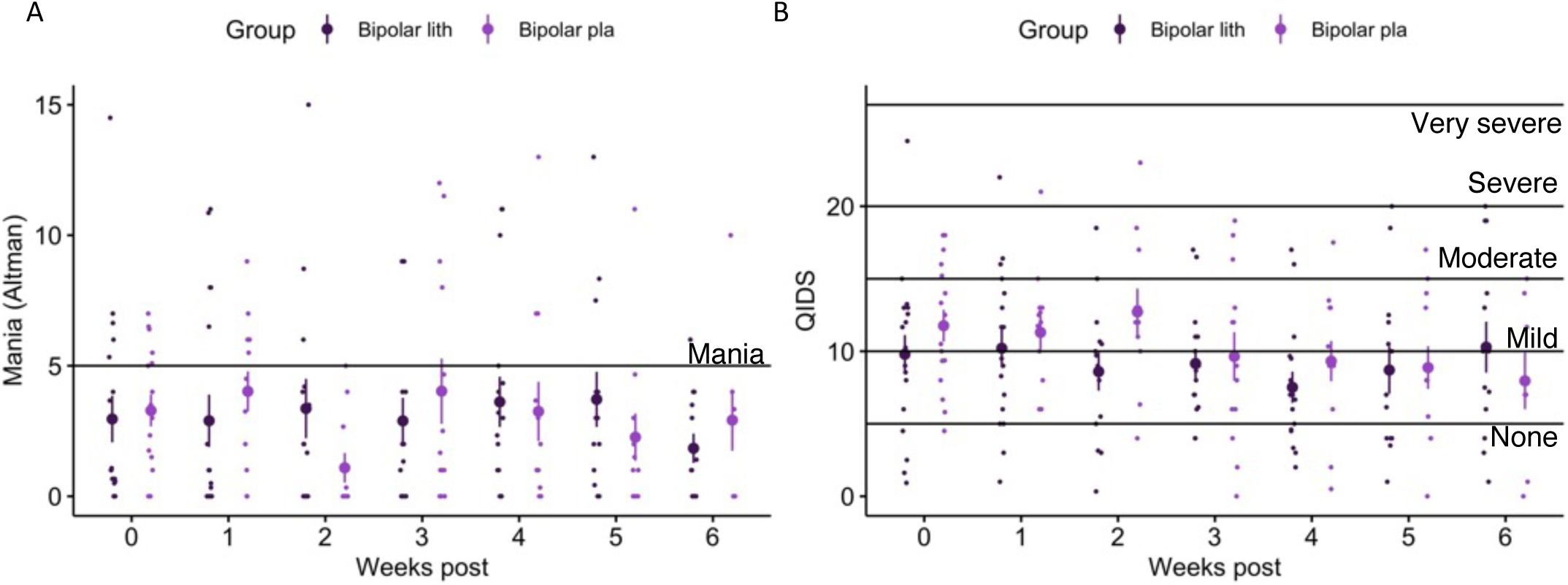
Mania (Altman Self Rating Mania Scale, A) and depression (Quick Inventory of Depressive Symptomatology, B) over the course of the study in patients with BD. Week zero is the average value of all weeks pre randomization (i.e. baseline) to lithium (black) vs. placebo. (purple) Horizontal lines show standard cut-offs. There were no significant differences between the groups (result of a regression predicting Altman or QIDS based on group, time and group*time, controlling for age and gender: group*time interaction: Altman 0.018, 95% CI [-0.10; 0.14]; QIDS: 0.05, 95% CI: [-0.1; 0.2]; main effect of group: Altman -0.70, 95% CI [-3.5; 2.0]; QIDS: -0.09, 95% CI: [-0.10; 0.02]; main effect of time: Altman: -0.01, 95% CI [-0.10; 0.07]; QIDS: -0.09, 95% CI: [-0.19; 0.02]).

**Figure S4.**
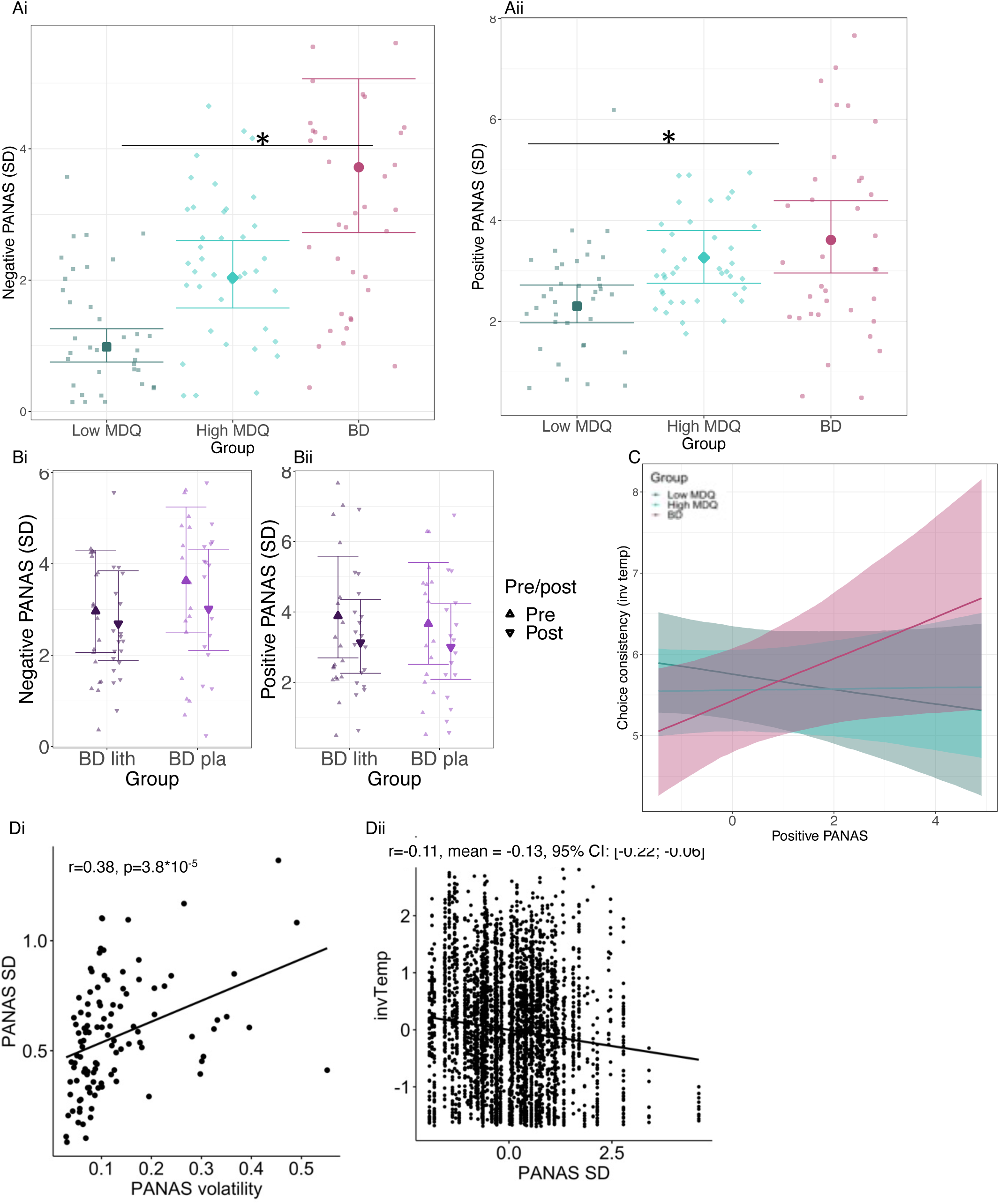
Mood (PANAS) variability (standard deviation). A and B as previously reported in Panchal et al. (7), shown here for ease of accessibility together with results relating mood to behaviour. A) BD gradient (ordered factor with low MDQ < high MDQ < patients with BD pre assignment to placebo or lithium) is linked to increased variability (standard deviation) for both positive and negative mood (PANAS). B) In contrast, lithium (as interaction term drug (lithium/placebo) * time (pre/post) does not affect variability of mood. See Table S8 for statistical values. C) Linking daily ratings of positive PANAS to inverse temperature (choice consistency) revealed an interaction with group (see Table S8). D) We adapted a more comprehensive model of mood variation from Pulcu et al. (1) (see supplementary methods [4] Bayesian mood instability models), fitting both mood variability and the links of mood variability to behaviour. Di) In the model, we captured separately the noisiness of mood ratings (PANAS SD, standard deviation) and the change in the average underlying moods (PANAS volatility). Dii) The groups differed in both measures of mood instability. Group was not included in the models. To test how predictive mood instability was for group, we trained regression models for out-of-sample leave-one-out predictions (see supplementary methods [4]). We found that for a model trained to predict low vs high MDQ group, % correct classification prediction was 72% (chance: 46%). When training to predict the three groups, classification was 61% correct (either when trained on all participants or omitting those that had a diagnosis of BD in the high MDQ group; chance: 32% correct). For the high MDQ participants with a BD diagnoses, 0% were misclassified as BD. Diii) The higher the mood variability (PANAS SD), the less consistent participants’ choices (i.e. lower inverse Temperature), mean =- 0.07, 95%CI [-0.15; -0.002]. We illustrate here the data across all measurements, but statistics were done in the full model taking the hierarchical structure of the data (i.e. several days per participant) into account. For other parameters, no consistent results emerged (i.e. changing the modelling approach slightly to capture mood instability and its link to behaviour in separate models meant that some results disappeared that were significant in the full model, suggesting that they were at least less reliable).

**Figure S5.**
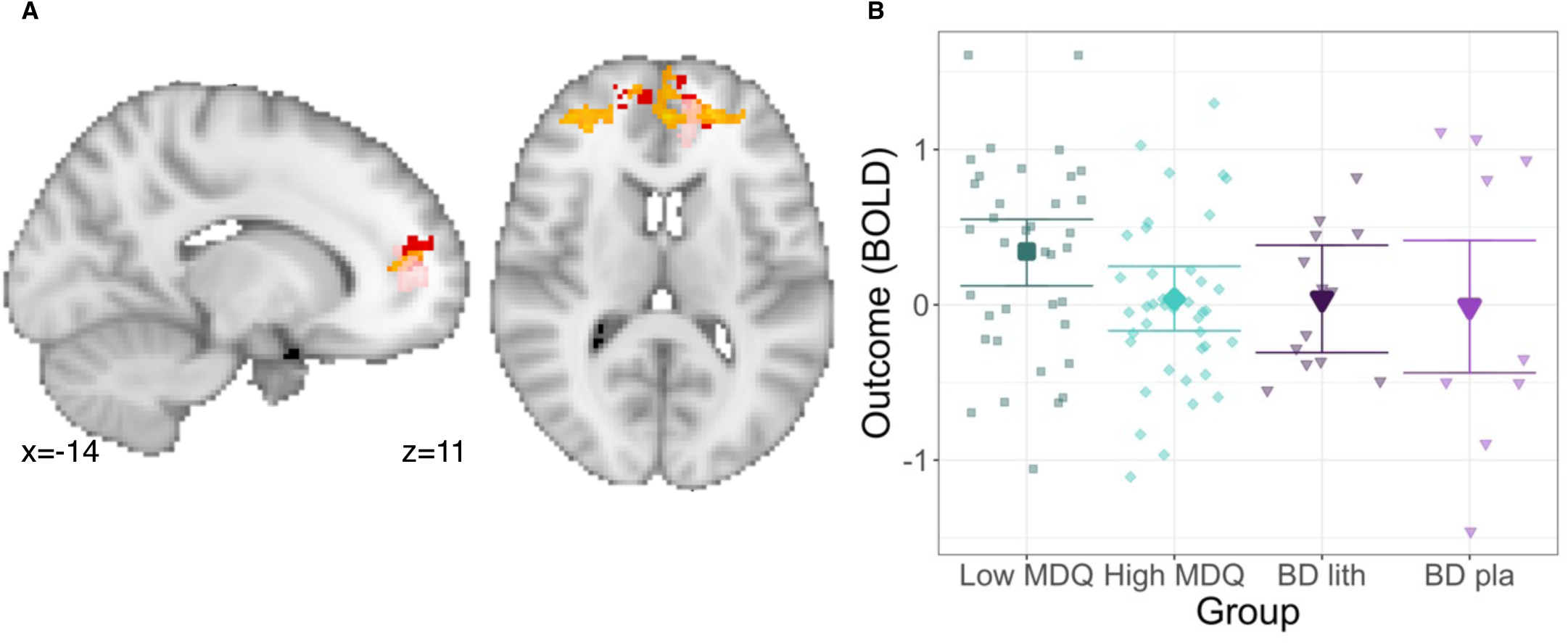
Whole-brain individual difference analyses. A) To complement Figure 4Aiii, we repeated the analyses whole brain, linking individual differences in the behaviour (at home) for the outcome history effect to neural signals for the last trial’s outcome at choice (orange) and for the reward/loss outcome signal (pink). Significant (p<0.05) whole-brain clusters overlapped with the area of the group difference in the last trial’s outcome signal at choice between the high and low MDQ groups (red, see Figure 4Ai). In particular, the result for the current trial outcome is noteworthy as that activation did not show any group differences. Together, this highlights a role for this medial frontal polar area in the outcome history effects. B) Despite the absence of whole-brain corrected differences between the high and low MDQ group for the outcome reward/loss activations, we also tested for group differences in the region of group differences for the last trial’s outcome at the time of the choice (Figure 4A, red activation in A). We found that in fact, the low MDQ group shows a greater outcome-related activation than the high MDQ group (mean = 0.5, 95% Bayesian CI: [0.06; 0.93]). There were no differences between the BD lithium and BD placebo groups (mean=-0.1, 95% CI: [-0.92; 0.76]). Participant numbers: n=99 (all available participants across the four groups).

**Table S1.** Parameter recovery, longitudinal data. (related to main text methods ‘Computational models’). Parametric (Pearson) correlations between ground-truth (‘t.’) and fitted (‘f.’) parameters. We simulated 400 participants with mean (‘_mean’) and standard (‘_sd’) from which choices for individual sessions of 20 trials were then generated (see supplementary methods [2A]). Simulated participants provided 47-50 sessions of data (50%-100% range of participants). Parameters: inverse temperature (β), sensitivity to loss utility (λ) and change in loss sensitivity after prev. trial win vs. loss (γ). Results show that recovery for mean parameters was very good (correlations between true and corresponding fitted, all >0.68). However, recovery for standard deviations was poor (e.g. γ_sd: r=0.24). Given this, we decided not to analyse group differences in standard deviations. Neither did we therefore attempt more complex models of variations of parameters across time, such as a volatility model (1).

| | $t.\beta\_mean$ | $t.\lambda\_mean$ | $t.\gamma\_mean$ | $t.\beta\_sd$ | $t.\lambda\_sd$ | $t.\gamma\_sd$ | $f.\beta\_mean$ | $f.\lambda\_mean$ | $f.\gamma\_mean$ | $f.\beta\_sd$ | $f.\lambda\_sd$ | $f.\gamma\_sd$ |
| --- | --- | --- | --- | --- | --- | --- | --- | --- | --- | --- | --- | --- |
| $t.\beta\_mean$ | | 0.018 | -0.095 | 0.150** | 0.024 | -0.077 | 0.875*** | -0.129** | -0.088 | -0.213*** | 0.204*** | 0.008 |
| $t.\lambda\_mean$ | 0.018 | | 0.064 | -0.000 | -0.055 | 0.024 | -0.307*** | 0.962*** | 0.060 | 0.318*** | 0.427*** | 0.592*** |
| $t.\gamma\_mean$ | -0.095 | 0.064 | | -0.045 | -0.045 | -0.042 | -0.103* | 0.061 | 0.911*** | 0.063 | -0.045 | 0.059 |
| $t.\beta\_sd$ | 0.150** | -0.000 | -0.045 | | -0.078 | -0.026 | 0.107* | -0.023 | -0.055 | 0.178*** | -0.037 | -0.011 |
| $t.\lambda\_sd$ | 0.024 | -0.055 | -0.045 | -0.078 | | -0.003 | 0.078 | -0.041 | -0.055 | 0.047 | 0.641*** | -0.099* |
| $t.\gamma\_sd$ | -0.077 | 0.024 | -0.042 | -0.026 | -0.003 | | -0.083 | 0.039 | -0.004 | -0.027 | -0.027 | 0.428*** |
| $f.\beta\_mean$ | 0.875*** | -0.307*** | -0.103* | 0.107* | 0.078 | -0.083 | | -0.423*** | -0.094 | -0.232*** | 0.057 | -0.212*** |
| $f.\lambda\_mean$ | -0.129** | 0.962*** | 0.061 | -0.023 | -0.041 | 0.039 | -0.423*** | | 0.055 | 0.379*** | 0.393*** | 0.575*** |
| $f.\gamma\_mean$ | -0.088 | 0.060 | 0.911*** | -0.055 | -0.055 | -0.004 | -0.094 | 0.055 | | 0.090 | -0.064 | 0.069 |
| $f.\beta\_sd$ | -0.213*** | 0.318*** | 0.063 | 0.178*** | 0.047 | -0.027 | -0.232*** | 0.379*** | 0.090 | | 0.198*** | 0.177*** |
| $f.\lambda\_sd$ | 0.204*** | 0.427*** | -0.045 | -0.037 | 0.641*** | -0.027 | 0.057 | 0.393*** | -0.064 | 0.198*** | | 0.216*** |
| $f.\gamma\_sd$ | 0.008 | 0.592*** | 0.059 | -0.011 | -0.099* | 0.428*** | -0.212*** | 0.575*** | 0.069 | 0.177*** | 0.216*** | |

Computed correlation used pearson-method with listwise-deletion.

**Table S2.** Model comparison with Akaishi Information Criterion (AIC) (related to main text methods ‘Computational models’). We compared three types of models. M1 (model used in main manuscript): decision variables as weighted combination of reward and loss (methods in main text); M2: decision variable containing exponential scaling of reward and loss magnitudes (supplementary methods [2B]); M3: weighted combination expected value, variance and skew (supplementary methods [2B]). Each of these models was run with different parameters being included or excluded (‘x’ in the table indicates inclusion) – parameter abbreviations: inv. temp = inverse temperature; outcome hist = outcome history; reward dist.= exponential distortion of reward magnitude; loss dist. = exponential distortion of loss magnitude. Annotations: ^1^Model of type M2, with outcome history included as linear weighing of the loss expected value; ^2^Model of type M2, with outcome history included in the exponential weighing; ^3^Model of type M2, with a shared parameter for exponential distorting of reward and loss magnitudes. AIC values are shown relative to the best fitting model, higher numbers indicate worse fit. Importantly, for all types of models, there is a model including outcome history that provides the best fit. While the model presented in the paper (M1) showed not the best fit to the data, it was retained it for ease of meaning of parameters (e.g. sensitivity to potential losses vs. sensitivity to skew) and analogy to previous decision making studies. Potentially, in the present study, the reason that a model with variance and skew provides a better fit is because total expected value of the two options was often very similar (Figure S1). However, we note, that the key behavioural findings of the paper, i.e. decrease in the outcome history effect with mood elevation gradient and decrease in loss aversion in BP are also captured in the both models M2 and M3, see Table S5. Though note that for the outcome history effect in M3, the 95%CI for the effect just about included zero [-0.0006;0.06], while the difference between the BD and low MDQ and high MDQ groups was significant). Parameter recovery for M2 and M3 type models in Table S3.

| Model type | inv. temp | variance | skew | outcome hist | reward dist. | loss dist. | loss weight | AIC |
| --- | --- | --- | --- | --- | --- | --- | --- | --- |
| M3 | x | x | x | x |  |  |  | 0 |
| M3 | x | x | x |  |  |  |  | 99 |
| M3 | x | x | x |  |  |  |  | 133 |
| M3 | x | x | x |  |  |  |  | 215 |
| M2 | x |  |  | x<sup>1</sup> | x | x |  | 3972 |
| M2 | x |  |  |  | x | x |  | 4052 |
| M2 | x |  |  | x<sup>2</sup> | x | x |  | 4131 |
| M3 | x |  | x |  |  |  |  | 5029 |
| M2 | x |  |  |  | x<sup>3</sup> | x<sup>3</sup> |  | 6126 |
| M1 | x |  |  | x |  |  | x | 6869 |
| M1 | x |  |  |  |  |  | x | 6875 |
| M2 | x |  |  |  |  | x |  | 6972 |
| M2 | x |  |  |  | x |  |  | 7127 |
| M3 | x | x |  |  |  |  |  | 7412 |
| M1/2/3 | x |  |  |  |  |  |  | 11431 |

**Table S3.** Parameter recovery for alternative models. (M2, M3, see Table S2 and supplementary methods [2B], related to main text methods ‘Computational models’). Shown are only the parameter recovery results for participant-wise means, the standard deviations showed equally bad recovery as for M1 (Table S1).

| **M3** | t.invTemp | t.var weight | t.skew weight | t.outcome history | f.invTemp | f.var weight | f.skew weight | f.outcome history |
| --- | --- | --- | --- | --- | --- | --- | --- | --- |
| t.invTemp |  | -0.058 | -0.109* | -0.08 | 0.867*** | -0.081 | -0.145** | -0.131** |
| t.var weight | -0.058 |  | 0.027 | 0.026 | -0.069 | 0.978*** | 0.01 | 0.049 |
| t.skew weight | -0.109* | 0.027 |  | 0.067 | -0.196*** | 0.015 | 0.977*** | 0.106* |
| t.outcome history | -0.08 | 0.026 | 0.067 |  | -0.082 | 0.017 | 0.058 | 0.907*** |
| f.invTemp | 0.867*** | -0.069 | -0.196*** | -0.082 |  | -0.081 | -0.220*** | -0.114* |
| f.var weight | -0.081 | 0.978*** | 0.015 | 0.017 | -0.081 |  | 0.001 | 0.041 |
| f.skew weight | -0.145** | 0.01 | 0.977*** | 0.058 | -0.220*** | 0.001 |  | 0.098 |
| f.outcome history | -0.131** | 0.049 | 0.106* | 0.907*** | -0.114* | 0.041 | 0.098 |  |
| **M2** | t.invTemp | t.loss mag dist | t.rew mag dist | t.outcome history | f.invTemp | f.loss mag dist | f.rew mag dist | f.outcome history |
| t.invTemp |  | 0.075 | -0.046 | -0.052 | 0.901*** | 0.099* | -0.091 | 0.006 |
| t.loss mag dist | 0.075 |  | -0.029 | -0.023 | 0.074 | 0.820*** | 0.112* | -0.009 |
| t.rew mag dist | -0.046 | -0.029 |  | 0.06 | -0.057 | 0.312*** | 0.905*** | 0.082 |
| t.outcome history | -0.052 | -0.023 | 0.06 |  | -0.06 | 0.019 | 0.071 | 0.914*** |
| f.invTemp | 0.901*** | 0.074 | -0.057 | -0.06 |  | 0.077 | -0.135** | 0.011 |
| f.loss mag dist | 0.099* | 0.820*** | 0.312*** | 0.019 | 0.077 |  | 0.423*** | 0.046 |
| f.rew mag dist | -0.091 | 0.112* | 0.905*** | 0.071 | -0.135** | 0.423*** |  | 0.09 |
| f.outcome history | 0.006 | -0.009 | 0.082 | 0.914*** | 0.011 | 0.046 | 0.09 |  |

**Table S4.** Computational modelling results. (related to Figure 2). Computational model parameters for the longitudinal data. A) Comparison of the three groups (mood elevation gradient, ordered factors across low MDQ, high MDQ, patients with BD, at baseline, i.e. pre-randomization to lithium or placebo). The groups differed in their loss sensitivity (patients with BD being least sensitive to losses) and outcome history effects (patients with BD being least sensitive to past trial outcomes). How participants performed the task changed over time (effect of ‘Day’), in particular they became more random (lower inverse temperature) and less sensitive to potential losses (loss sensitivity). When repeating the analyses, but omitting the 5 participants from the high MDQ group that were given a BD diagnosis (Table 1) during the intake interview, results remained broadly the same. For loss sensitivity, we find the same results as before (-0.26; 95% CI: [- 0.48; -0.03]). For the outcome history effect, the group effect is not quite significant anymore, but numerically very close to the previous finding (-0.05; 95%CI: [-0.11; 0.0006]). When repeating the analyses and including as additional regressor for each session the time (days) since the previous session, we found numerically very similar results (Loss sensitivity: -0.25; 95%CI: [-0.47;-0.03]; outcome history: -0.0586, 95%CI: [-0.105; 0.0026]). Repeating the analyses and replacing group assignment by the Altman Mania score (‘Mania’, continuous measure, available for all but one participant in the BD group [later assigned to lithium] and 3 in the high MDQ group), outcome history remains significant, while loss sensitivity is no longer significant (though trend in the same direction as considering group). B) Comparison for the effect of lithium vs. placebo in hierarchical models (Main text Methods, section ‘Model fitting’, term of interest is the interaction drug (lithium/placebo) * time (pre/post)). No significant group differences were found. Values are means and 95% Bayesian Credible Intervals; for comparisons between groups, significance is defined as 95% intervals not including zero. All estimates were obtained from hierarchical regression models (Main text Methods, section ‘Model fitting’).

| A) Low MDQ, High MDQ, Bipolar disorder (BD) groups (baseline) |  |  |  |
| --- | --- | --- | --- |
| Group | Choice consist. ($\beta$) | Loss sensitivity ($\lambda$) | Outcome history effect ($\gamma$) |
| 3 group gradient | -0.154 [-0.5698 0.2498] | -0.269 [-0.4868 -0.0519] | -0.053 [-0.1104 -3e-04] |
| High vs low MDQ | -0.42 [-1.15 0.36] | -0.01 [-0.41 0.39] | -0.05 [-0.11 0.03] |
| BD vs high MDQ | 0.14 [-0.62 0.9] | -0.5 [-0.93 -0.08] | -0.07 [-0.19 0.05] |
| BD vs low MDQ | -0.3 [-1 0.52] | -0.52 [-0.94 -0.08] | -0.12 [-0.24 0] |
| BD | 5.84 [5.24 6.49] | 0.89 [0.56 1.23] | 0 [-0.11 0.1] |
| High MDQ | 5.71 [5.13 6.29] | 1.4 [1.12 1.69] | 0.07 [0.02 0.12] |
| Low MDQ | 6.13 [5.55 6.71] | 1.41 [1.13 1.72] | 0.11 [0.06 0.16] |
| Day | -0.555 [-0.763 -0.339] | -0.239 [-0.329 -0.155] | -0.03 [-0.062 0.002] |
| Mania | -0.208 [-0.4911 0.0728] | -0.087 [-0.2375 0.0625] | -0.039 [-0.072 -0.0056] |
| B) BD groups, pre/post * lithium/placebo |  |  |  |
| Group | Choice consist. ($\beta$) | Loss sensitivity ($\lambda$) | Outcome history effect ($\gamma$) |
| Lith/pla x pre/post | 0.193 [-0.9163 1.2654] | -0.01 [-0.5146 0.4884] | -0.11 [-0.2983 0.0797] |
| Lith pre vs post | 0.36 [-0.49 1.2] | -0.18 [-0.53 0.16] | -0.06 [-0.19 0.08] |
| Pla pre vs post | 0.17 [-0.78 1.09] | -0.17 [-0.54 0.21] | 0.05 [-0.09 0.2] |
| Lith vs pla (pre) | 0.76 [-0.42 1.83] | 0.19 [-0.25 0.61] | -0.07 [-0.23 0.1] |
| Lith vs pla (post) | 0.55 [-0.6 1.73] | 0.2 [-0.22 0.62] | 0.04 [-0.08 0.17] |
| Lith (pre) | 6.36 [5.43 7.29] | 1.09 [0.8 1.39] | -0.03 [-0.15 0.09] |
| Placebo (pre) | 5.61 [4.61 6.6] | 0.9 [0.58 1.23] | 0.04 [-0.08 0.17] |
| Lith (post) | 6 [5.19 6.88] | 1.26 [0.97 1.55] | 0.03 [-0.06 0.11] |
| Placebo (post) | 5.44 [4.5 6.35] | 1.06 [0.76 1.38] | -0.01 [-0.1 0.09] |

**Table S5.** Group comparisons for alternative models. (M2, M3, see Table S2 and supplementary methods [2B]). A) In M3, instead of considering reward and loss expected values/ utility separately, we consider total expected value, variance and skew. Now, the previous (M1) decreased loss risk aversion with BD expressed itself as increased preference for options with higher variance (see Figure S1 for relationship – high correlation- between risk of loss and variance). The mood elevation gradient is again linked to decreased adaptation across trials, here captured as decreased preference to options with high variance after win on previous trial (in M1: decreased avoidance of risk of losing). B) In M2, the model has parameters for the (exponential) distortion of reward magnitudes and loss magnitudes. Outcome history effects are captured as linear weighting of the loss expected value. Again, group differences captured are conceptually very similar to M1. As we could not fit a model (due to low trial numbers per session) including both the linear and exponential effects of loss sensitivity, future studies will need to be done to describe the specific shape of the increased loss sensitivity more precisely. Values are means and 95% Bayesian Credible Intervals; for comparisons between groups, significance is defined as 95% intervals not including zero.

| **A) M3: Expected value, variance, skew** |  |  |  |  |
| --- | --- | --- | --- | --- |
| **Ai) M3: Low MDQ, High MDQ, Bipolar disorder (BD, baseline) groups** |  |  |  |  |
| Group | Choice consist. ($\beta$) | Var Weight | Skew Weight | Outcome history effect |
| 3 group gradient | -0.114 [-0.4411 0.2132] | 0.108 [0.0072 0.2066] | 0.047 [-0.0497 0.145] | 0.027 [-6e-04 0.0556] |
| High vs low MDQ | 0.05 [-0.53 0.62] | 0.01 [-0.17 0.19] | 0.04 [-0.13 0.22] | 0.01 [-0.03 0.04] |
| BD vs high MDQ | -0.26 [-0.82 0.42] | 0.21 [0.02 0.42] | 0.06 [-0.13 0.27] | 0.06 [0 0.11] |
| BD vs low MDQ | -0.21 [-0.81 0.42] | 0.22 [0.02 0.42] | 0.1 [-0.09 0.31] | 0.06 [0.01 0.12] |
| BD | 2.62 [2.12 3.13] | 0.23 [0.07 0.38] | 0.1 [-0.05 0.26] | 0.01 [-0.04 0.06] |
| High MDQ | 2.87 [2.43 3.33] | 0.02 [-0.12 0.14] | 0.04 [-0.08 0.17] | -0.04 [-0.07 -0.02] |
| Low MDQ | 2.83 [2.36 3.27] | 0.01 [-0.12 0.14] | 0 [-0.14 0.13] | -0.05 [-0.08 -0.02] |
| Day | -0.133 [-0.297 0.033] | 0.047 [0.015 0.08] | 0.108 [0.07 0.147] | 0.014 [-0.001 0.029] |
| **Aii) M3: BD groups, pre/post \* lithium/placebo** |  |  |  |  |
| Group | Choice consist. ($\beta$) | Var Weight | Skew Weight | Outcome history effect |
| Lith/pla x pre/post | -0.273 [-1.1636 0.6362] | 0.152 [-0.0605 0.364] | -0.217 [-0.4738 0.0465] | -0.006 [-0.1042 0.0895] |
| Lith pre vs post | -0.28 [-0.96 0.4] | 0.16 [0.02 0.32] | -0.04 [-0.22 0.14] | 0.02 [-0.05 0.09] |
| Pla pre vs post | -0.01 [-0.72 0.7] | 0.01 [-0.14 0.18] | 0.18 [-0.03 0.37] | 0.02 [-0.05 0.1] |
| Lith vs pla (pre) | 0.61 [-0.23 1.5] | 0.09 [-0.12 0.3] | -0.11 [-0.39 0.16] | 0 [-0.09 0.08] |
| Lith vs pla (post) | 0.89 [-0.03 1.81] | -0.06 [-0.25 0.12] | 0.1 [-0.2 0.43] | 0 [-0.06 0.06] |
| Lith (pre) | 2.83 [2.14 3.49] | 0.24 [0.1 0.39] | -0.03 [-0.22 0.16] | 0.01 [-0.05 0.07] |
| Placebo (pre) | 2.22 [1.53 2.92] | 0.15 [-0.01 0.3] | 0.09 [-0.11 0.28] | 0.02 [-0.05 0.08] |
| Lith (post) | 3.11 [2.45 3.77] | 0.08 [-0.05 0.2] | 0.01 [-0.19 0.22] | -0.01 [-0.05 0.03] |
| Placebo (post) | 2.22 [1.54 2.93] | 0.14 [0 0.27] | -0.09 [-0.32 0.14] | -0.01 [-0.05 0.04] |
| **B) M2: exponential distortions of magnitudes** |  |  |  |  |
| **Bi) M2: Low MDQ, High MDQ, BD (baseline) groups** |  |  |  |  |
| Group | Choice consist. ($\beta$) | Rew mag dist | Loss mag dist | Outcome history effect |
| 3 group gradient | -0.362 [-0.8268 0.1023] | 0.046 [-0.0291 0.1216] | -0.078 [-0.1551 -1e-04] | 0.023 [3e-04 0.0532] |
| High vs low MDQ | -0.12 [-1 0.76] | 0.04 [-0.1 0.17] | 0 [-0.13 0.13] | 0.02 [0 0.04] |
| BD vs high MDQ | -0.48 [-1.3 0.43] | 0.06 [-0.1 0.21] | -0.16 [-0.32 -0.02] | 0.03 [-0.04 0.1] |
| BD vs low MDQ | -0.6 [-1.48 0.25] | 0.1 [-0.06 0.26] | -0.16 [-0.31 -0.01] | 0.05 [-0.02 0.12] |
| BD | 7.12 [6.4 7.9] | 1.34 [1.22 1.47] | 1.32 [1.2 1.44] | 0.04 [-0.03 0.1] |
| High MDQ | 7.6 [6.91 8.34] | 1.29 [1.19 1.39] | 1.49 [1.39 1.58] | 0.01 [-0.01 0.03] |
| Low MDQ | 7.73 [7.01 8.47] | 1.25 [1.15 1.34] | 1.48 [1.39 1.57] | -0.01 [-0.03 0.01] |
| Day | -0.668 [-0.928 -0.409] | 0.056 [0.027 0.085] | 0.029 [-0.002 0.06] | 0.012 [0.001 0.023] |
| **Bii) M2: BD groups, pre/post \* lithium/placebo** |  |  |  |  |
| Group | Choice consist. ($\beta$) | Rew mag dist | Loss mag dist | Outcome history effect |
| Lith/pla x pre/post | -0.073 [-1.2749 1.123] | -0.085 [-0.2576 0.0919] | -0.202 [-0.4042 3e-04] | 0.054 [-0.1137 0.2272] |
| Lith pre vs post | -0.16 [-1.05 0.74] | 0.01 [-0.11 0.12] | -0.07 [-0.21 0.08] | -0.01 [-0.12 0.11] |
| Pla pre vs post | -0.09 [-1.15 0.99] | 0.09 [-0.03 0.23] | 0.13 [-0.02 0.29] | -0.06 [-0.2 0.07] |
| Lith vs pla (pre) | 0.83 [-0.4 2.01] | -0.02 [-0.18 0.15] | -0.09 [-0.36 0.2] | 0.04 [-0.11 0.18] |
| Lith vs pla (post) | 0.9 [-0.5 2.29] | 0.07 [-0.13 0.27] | 0.11 [-0.14 0.35] | -0.02 [-0.13 0.09] |
| Lith (pre) | 7.55 [6.52 8.57] | 1.29 [1.17 1.4] | 1.26 [1.07 1.46] | 0.02 [-0.08 0.12] |
| Placebo (pre) | 6.71 [5.62 7.8] | 1.3 [1.18 1.42] | 1.36 [1.16 1.56] | -0.02 [-0.12 0.1] |
| Lith (post) | 7.71 [6.64 8.76] | 1.28 [1.15 1.42] | 1.33 [1.17 1.5] | 0.03 [-0.04 0.09] |
| Placebo (post) | 6.81 [5.63 7.94] | 1.21 [1.06 1.35] | 1.22 [1.05 1.4] | 0.05 [-0.04 0.12] |

**Table S6.** FMRI session computational parameters. (related to Figure 2). Values are reported separately for each of the four groups (Bipolar participants on lithium (‘Bip Lith’), bipolar participants on placebo (‘Bip Pla’), healthy volunteers with low or high mood instability (‘Low MDQ’, ‘High MDQ’) as means and 95% Bayesian Credible Intervals (intervals not including zero are significant). All estimates were obtained from linear regression models allowing correcting for age and gender. Group differences were computed across all four groups (‘4 group diff’). Group differences are also reported separately comparing high and low mood instability participants (‘High vs. low MDQ’) and lithium vs. placebo participants (‘Lith vs pla’). There were no significant group differences.

| Group | Choice consist. ($\beta$) | Loss sensitivity ($\lambda$) | Outcome history effect ($\gamma$) |
| --- | --- | --- | --- |
| Lith (post) | 8.8 [7.08 10.52] | 1.48 [0.92 2.03] | -0.07 [-0.31 0.17] |
| Pla (post) | 6.58 [4.55 8.48] | 1.58 [0.95 2.29] | -0.01 [-0.27 0.27] |
| High MDQ | 7.06 [6.08 8.08] | 1.76 [1.38 2.07] | 0.13 [-0.02 0.27] |
| Low MDQ | 7.34 [6.28 8.34] | 1.73 [1.38 2.09] | 0.12 [-0.03 0.25] |
| 4 Group diff (post) | -0.002 [-0.74 0.67] | -0.059 [-0.278 0.157] | -0.051 [-0.141 0.039] |
| Lith vs pla (post) | 2.23 [-0.3 4.95] | -0.1 [-0.96 0.75] | -0.06 [-0.42 0.28] |
| High vs low MDQ | -0.28 [-1.61 1.21] | 0.03 [-0.46 0.53] | 0.02 [-0.17 0.21] |

**Table S7.**
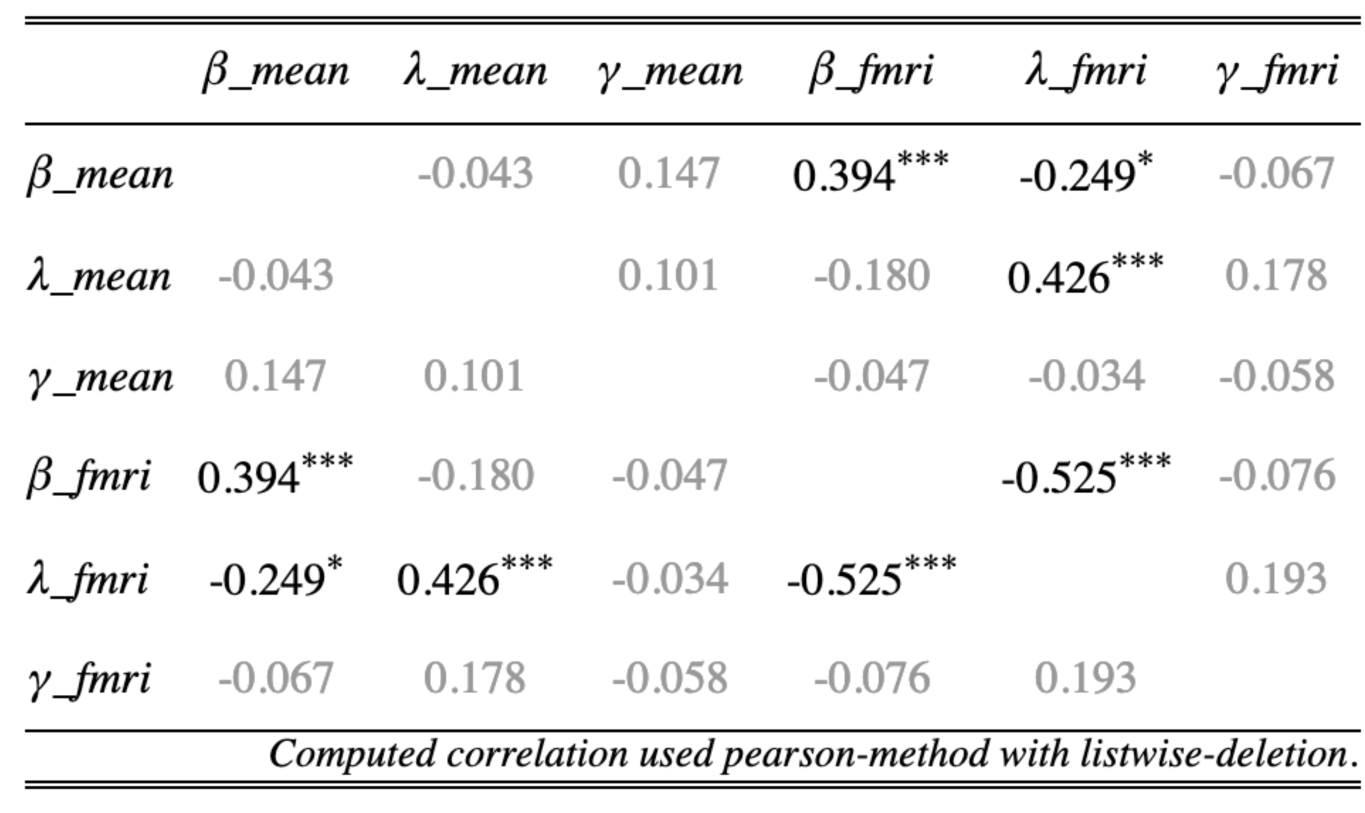
Correlations between parameters from longitudinal and FMRI data. (related to Figure 2). For all but one parameter, computational parameters derived from longitudinal measurements and those obtained during the FMRI scan correlate significantly. Only for the outcome history effect parameter (γ) are the correlations not significant. Correlations were computed using Pearson correlations across combined data from all four participant groups.

| | $\beta_{mean}$ | $\lambda_{mean}$ | $\gamma_{mean}$ | $\beta_{fmri}$ | $\lambda_{fmri}$ | $\gamma_{fmri}$ |
| --- | --- | --- | --- | --- | --- | --- |
| $\beta_{mean}$ | | -0.043 | 0.147 | 0.394*** | -0.249* | -0.067 |
| $\lambda_{mean}$ | -0.043 | | 0.101 | -0.180 | 0.426*** | 0.178 |
| $\gamma_{mean}$ | 0.147 | 0.101 | | -0.047 | -0.034 | -0.058 |
| $\beta_{fmri}$ | 0.394*** | -0.180 | -0.047 | | -0.525*** | -0.076 |
| $\lambda_{fmri}$ | -0.249* | 0.426*** | -0.034 | -0.525*** | | 0.193 |
| $\gamma_{fmri}$ | -0.067 | 0.178 | -0.058 | -0.076 | 0.193 | |
| *Computed correlation used pearson-method with listwise-deletion.* |  |  |  |  |  |  |

**Table S8.** Group difference for mood (PANAS) mean and standard deviations and impact of task outcomes on momentary mood (VAS). A) Comparison of the three groups (mood elevation gradient, ordered factors across low MDQ, high MDQ, patients with BD [i.e. pre randomization to lithium or placebo]). The groups differed in variability (standard deviation (log scale) for positive and negative PANAS with patients with BD showing the highest variability. Groups also differed in the mean values for negative PANAS. B) Comparison for the effect of lithium vs. placebo in hierarchical models (Main text Methods, section ‘Model fitting’, term of interest is the interaction drug (lithium/placebo) * time (pre i.e. baseline /post)). No significant group differences were found. C) In a regression predicting changes in mood rated on a visual analogue scale (VAS) post completing the daily task vs. pre, there was an overall effect that the higher the total reward, and the lower the total loss (i.e. more positive number), the more mood improves. However, this impact of task outcomes on mood was not affected by the mood elevation gradient (‘3 Group x outcome’). Values are means and 95% Bayesian Credible Intervals; significance is defined as 95% intervals not including zero. All estimates were obtained from hierarchical regression models (Main text Methods, section ‘Model fitting’). D) In separate regressions, we assessed the impact of mood behaviour the daily tasks on behaviour. Mood was measured as positive or negative PANAS or as the total PANAS (positive minus negative PANAS). The regressions were of the same form as throughout the paper (e.g. S4), additionally including mood and/or an interaction between mood and BD gradient. For completeness, we report the results here for regressions only including mood, not BD gradient (‘not ctr gr’), controlling BD gradient in addition to mood (‘ctr gr’), controlling for an interaction between mood and BD gradient (‘ctr interact’). We report also the interactions between mood and BD gradient (‘x 3gr gradient’) and the result for BD gradient, controlling intearctions with mood (‘ctr mood interact’). We find (figure S4C for illustration) that in the BD group, choice consistency is higher (i.e. less choice noisiness) when positive PANAS is higher. While there appears to be an impact of negative PANAS on outcome history, this only emerges when including the BD gradient as a regressor, making it difficult to interpret (one could speculate that a ‘masking’ effect is present because neg PANAS and group have the opposite impact on outcome history and neg PANAS differs between the groups).

| **A) Low MDQ, High MDQ, Bipolar disorder (BD) groups (baseline)** |  |  |  |  |
| --- | --- | --- | --- | --- |
| **Group** | **Positive PANAS (mean)** | **Negative PANAS (mean)** | **Positive PANAS (sd)** | **Negative PANAS (sd)** |
| 3 group gradient | 0.15 [-0.6434 0.9624] | 2.61 [1.9555 3.2551] | 0.221 [0.1118 0.3321] | 0.642 [0.4558 0.8305] |
| High - low MDQ | 0.16 [-1.29 1.67] | 1.26 [-0.02 2.47] | 0.35 [0.15 0.56] | 0.73 [0.41 1.06] |
| BD - high MDQ | 0.04 [-1.22 1.38] | 2.1 [0.9 3.25] | 0.1 [-0.12 0.34] | 0.6 [0.23 0.95] |
| BD - low MDQ | 0.2 [-1.11 1.52] | 3.36 [2.18 4.45] | 0.45 [0.22 0.68] | 1.33 [0.94 1.71] |
| BD | 6.76 [5.63 7.86] | 4.94 [3.97 5.87] | 1.15 [0.97 1.33] | 1.13 [0.84 1.41] |
| High MDQ | 6.73 [5.75 7.75] | 2.83 [2.03 3.73] | 1.05 [0.9 1.19] | 0.53 [0.3 0.75] |
| Low MDQ | 6.56 [5.41 7.69] | 1.57 [0.73 2.53] | 0.7 [0.56 0.86] | -0.2 [-0.43 0.05] |
| Day | -0.296 [-0.56 -0.049] | -0.007 [-0.139 0.119] | NA | NA |
| **B) BD groups, pre/post \* lithium/placebo** |  |  |  |  |
| **Group** | **Positive PANAS (mean)** | **Negative PANAS (mean)** | **Positive PANAS (sd)** | **Negative PANAS (sd)** |
| Lith/pla x pre/post | -0.274 [-1.5488 1.019] | 0.264 [-0.8541 1.3526] | 0.013 [-0.4098 0.4187] | -0.086 [-0.4867 0.3164] |
| Lith pre vs post | 1.01 [-0.03 2.08] | 0.86 [-0.01 1.72] | 0.22 [-0.06 0.51] | 0.1 [-0.19 0.36] |
| Pla pre vs post | 1.3 [0.12 2.58] | 0.58 [-0.41 1.59] | 0.2 [-0.11 0.5] | 0.18 [-0.12 0.46] |
| Lith vs pla (pre) | 0.2 [-1.61 1.86] | -0.35 [-2 1.31] | 0.06 [-0.43 0.49] | -0.2 [-0.65 0.25] |
| Lith vs pla (post) | 0.49 [-1.59 2.51] | -0.62 [-2.47 1.27] | 0.04 [-0.36 0.47] | -0.11 [-0.57 0.32] |
| Lith (pre) | 7.92 [5.98 9.87] | 6.19 [4.41 8.04] | 1.17 [0.84 1.48] | 0.88 [0.54 1.18] |
| Placebo (pre) | 7.72 [5.78 9.7] | 6.55 [4.62 8.35] | 1.11 [0.77 1.45] | 1.08 [0.75 1.39] |
| Lith (post) | 6.89 [4.98 8.9] | 5.33 [3.57 7.3] | 0.95 [0.66 1.23] | 0.78 [0.46 1.07] |
| Placebo (post) | 6.43 [4.48 8.6] | 5.97 [4.09 8.02] | 0.91 [0.59 1.19] | 0.89 [0.58 1.21] |
| **C) Group differences in impact of gambling outcomes on mood** |  |  |  |  |
| **Regressor** | **Total gain** |  | **Total loss** | **Total gain minus loss** |
| Outcome->Mood change | 0.843 [0.6303 1.0599] |  | 0.942 [0.7177 1.1755] | 1.026 [0.7737 1.286] |
| 3 group gradient | 0.12 [-0.0254 0.2764] |  | 0.188 [0.0291 0.3525] | 0.133 [-0.0037 0.2764] |
| 3 group gradient x outcome | -0.083 [-0.2753 0.0951] |  | -0.146 [-0.3453 0.0576] | -0.123 [-0.3475 0.0926] |
| **D) Impact of mood before the task on task behaviour** |  |  |  |  |
| **Regressor** | **Choice consist. ($\beta$)** | **Loss sensitivity ($\lambda$)** | **Outcome history ($\gamma$)** |  |
| totalPANAS (not ctr gr) | 0.034 [-0.0835 0.1533] | 0.007 [-0.0488 0.061] | -0.01 [-0.0396 0.0187] |  |
| totalPANAS (ctr gr) | 0.03 [-0.0856 0.1478] | 0 [-0.0548 0.053] | -0.017 [-0.0484 0.014] |  |
| totalPANAS (ctr interact) | -0.093 [-0.2803 0.086] | -0.007 [-0.0973 0.0834] | -0.022 [-0.0708 0.0239] |  |
| totalPANAS x 3gr gradient | 0.157 [-0.0078 0.3185] | 0.005 [-0.0666 0.0772] | 0.005 [-0.0355 0.0438] |  |
| 3 group gradient (ctr totalPANAS interact) | -0.095 [-0.5057 0.3175] | -0.268 [-0.4829 -0.0452] | -0.059 [-0.1189 -0.0041] |  |
| posPANAS (not ctr gr) | 0.03 [-0.0933 0.158] | -0.015 [-0.0719 0.0419] | 0 [-0.0299 0.0306] |  |
| posPANAS (ctr gr) | 0.009 [-0.0289 0.0491] | -0.009 [-0.0466 0.0287] | 0.002 [-0.0367 0.0407] |  |
| posPANAS (ctr interact) | -0.093 [-0.2756 0.0794] | -0.017 [-0.1056 0.0673] | -0.001 [-0.047 0.0439] |  |
| posPANAS x 3gr gradient | 0.177 [0.0059 0.3461] | 0.002 [-0.0732 0.0782] | 0.001 [-0.0417 0.0412] |  |
| 3 group gradient (ctr posPANAS interact) | -0.166 [-0.5529 0.2334] | -0.265 [-0.4898 -0.038] | -0.056 [-0.1126 -0.0032] |  |
| negPANAS (not ctr gr) | -0.031 [-0.1628 0.0978] | -0.037 [-0.0992 0.0274] | 0.019 [-0.0122 0.0505] |  |
| negPANAS (ctr gr) | -0.007 [-0.0487 0.0359] | -0.014 [-0.0576 0.0298] | 0.045 [0.0025 0.0872] |  |
| negPANAS (ctr interact) | 0.036 [-0.2153 0.2919] | -0.012 [-0.1293 0.1091] | 0.071 [0.0039 0.1487] |  |
| negPANAS x 3gr gradient | -0.053 [-0.2339 0.1254] | -0.008 [-0.0952 0.0775] | -0.027 [-0.0747 0.0178] |  |
| 3 group gradient (ctr negPANAS interact) | -0.114 [-0.5185 0.3209] | -0.232 [-0.4619 0.0057] | -0.071 [-0.1345 -0.0138] |  |

**Table S9.**
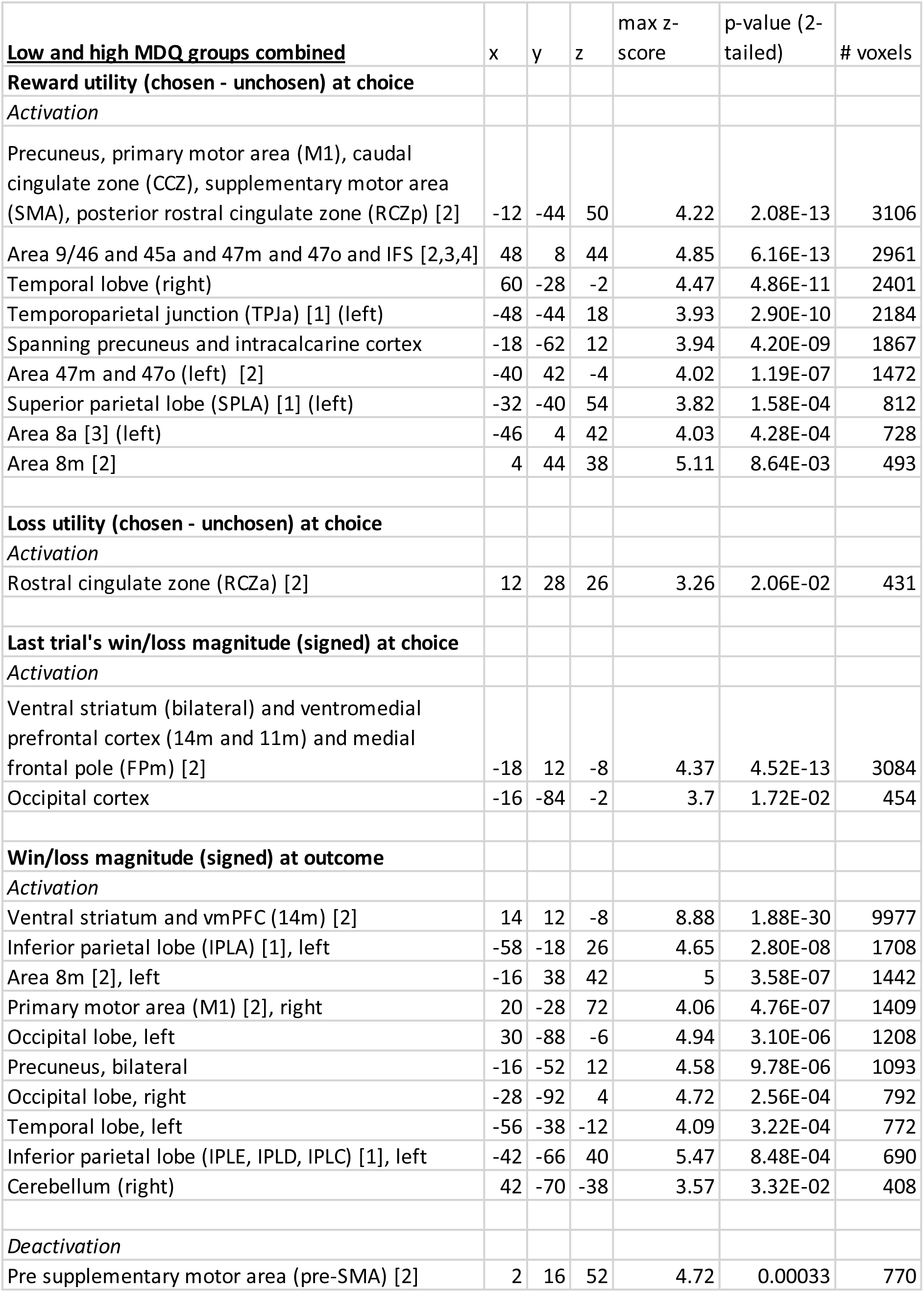
General task brain (de)activations. (related to Figure 3). Data across the low and high mood instability (MDQ) groups was combined to identify general brain (de)activations during the task. Coordinates are reported in MNI space. Significance was determined using cluster-based thresholding (methods section “FMRI analysis – whole-brain”), with inclusion threshold: z=2.3 and significance p<0.05 two-tailed. The maximum z-value of the cluster, the p-value and number of voxels are given for each cluster. Anatomical labels are based on: [1] (2)) [2] (3), [3] (4), [4] (5), [5] (6).

**Table S10.** Whole-brain group comparisons. (related to Figure 4). A) Comparisons of the low vs high mood elevation volunteers. Repeating the group comparisons in the ROI, but excluding participants from the high MDQ group who had a BD diagnosis (n=5), results were still significant: estimate = 1.00, 95%CI=[0.6; 1.39]. B) Comparisons of the patients with BD assigned to placebo or lithium. All cluster-based thresholded, inclusion threshold: z=2.3, significance p<0.05 two-tailed. The maximum z-value of the cluster, the p-value and number of voxels are given for each cluster. Anatomical labels are based on: [1] (2)) [2] (3), [3] (4), [4] (5), [5] (6).

| **A Low vs high MDQ groups** | x | y | z | max z-score | p-value (2-tailed) | # voxels |
| --- | --- | --- | --- | --- | --- | --- |
| **Last trial's win/loss magnitude (signed) at choice** |  |  |  |  |  |  |
| *Low > high MDQ* |  |  |  |  |  |  |
| Medial frontal pole (FPm), area 9m [2] | -10 | 56 | 16 | 3.52 | 0.0378 | 398 |
| **B Bipolar lithium vs. placebo groups - exploratory** |  |  |  |  |  |  |
| **Win/loss magnitude (signed) at outcome** |  |  |  |  |  |  |
| *Placebo > Lithium* |  |  |  |  |  |  |
| Dorsolateral prefrontal cortex (Area 46 [5]) and lateral frontal pole [2], Inferior frontal sulcus (IFS), (right) | 38 | 48 | 0 | 3.49 | 0.00898 | 503 |

## References

1. Kessing LV, Bauer M, Nolen WA, Severus E, Goodwin GM, Geddes J (2018): Effectiveness of maintenance therapy of lithium vs other mood stabilizers in monotherapy and in combinations: a systematic review of evidence from observational studies. Bipolar Disord 20: 419–431.

2. Bonsall MB, Wallace-Hadrill SMA, Geddes JR, Goodwin GM, Holmes EA (2012): Nonlinear time-series approaches in characterizing mood stability and mood instability in bipolar disorder. Proc R Soc B Biol Sci 279: 916–924.

3. Pulcu E, Saunders KEA, Harmer CJ, Harrison PJ, Goodwin GM, Geddes JR, Browning M (2022): Using a generative model of affect to characterize affective variability and its response to treatment in bipolar disorder. Proc Natl Acad Sci 119: e2202983119.

4. Eldar E, Niv Y (2015): Interaction between emotional state and learning underlies mood instability. Nat Commun 6: 6149.

5. Eldar E, Rutledge RB, Dolan RJ, Niv Y (2016): Mood as representation of momentum. Trends Cogn Sci 20: 15–24.

6. Mason L, Eldar E, Rutledge RB (2017): Mood instability and reward dysregulation—a neurocomputational model of bipolar disorder. JAMA Psychiatry 74: 1275–1276.

7. Moningka H, Mason L (2024): Misperceiving Momentum: Computational Mechanisms of Biased Striatal Reward Prediction Errors in Bipolar Disorder. Biol Psychiatry Glob Open Sci 4: 100330.

8. Taquet M, Quoidbach J, Montjoye Y-A de, Desseilles M, Gross JJ (2016): Hedonism and the choice of everyday activities. Proc Natl Acad Sci 113: 9769–9773.

9. Taquet M, Quoidbach J, Gross JJ, Saunders KEA, Goodwin GM (2020): Mood Homeostasis, Low Mood, and History of Depression in 2 Large Population Samples. JAMA Psychiatry. 10.1001/jamapsychiatry.2020.0588

10. Zhang K, Clark L (2020): Loss-chasing in gambling behaviour: neurocognitive and behavioural economic perspectives. Curr Opin Behav Sci 31: 1–7.

11. Diekhof EK, Kaps L, Falkai P, Gruber O (2012): The role of the human ventral striatum and the medial orbitofrontal cortex in the representation of reward magnitude – An activation likelihood estimation meta-analysis of neuroimaging studies of passive reward expectancy and outcome processing. Neuropsychologia 50: 1252–1266.

12. Sescousse G, Caldú X, Segura B, Dreher J-C (2013): Processing of primary and secondary rewards: A quantitative meta-analysis and review of human functional neuroimaging studies. Neurosci Biobehav Rev 37: 681–696.

13. Lopez-Gamundi P, Yao Y-W, Chong TT-J, Heekeren HR, Mas-Herrero E, Marco-Pallarés J (2021): The neural basis of effort valuation: A meta-analysis of functional magnetic resonance imaging studies. Neurosci Biobehav Rev 131: 1275–1287.

14. Scholl J, Kolling N, Nelissen N, Wittmann MK, Harmer CJ, Rushworth MF (2015): The good, the bad, and the irrelevant: neural mechanisms of learning real and hypothetical rewards and effort. J Neurosci 35: 11233–11251.

15. Brandl F, Le Houcq Corbi Z, Mulej Bratec S, Sorg C (2019): Cognitive reward control recruits medial and lateral frontal cortices, which are also involved in cognitive emotion regulation: A coordinate-based meta-analysis of fMRI studies. NeuroImage 200: 659–673.

16. Koban L, Lee S, Schelski DS, Simon M-C, Lerman C, Weber B, et al. (2023): An fMRI-Based Brain Marker of Individual Differences in Delay Discounting. J Neurosci 43: 1600– 1613.

17. Mason L, O’Sullivan N, Montaldi D, Bentall RP, El-Deredy W (2014): Decision-making and trait impulsivity in bipolar disorder are associated with reduced prefrontal regulation of striatal reward valuation. Brain 137: 2346–2355.

18. Whittaker JR, Foley SF, Ackling E, Murphy K, Caseras X (2018): The functional connectivity between the nucleus accumbens and the ventromedial prefrontal cortex as an endophenotype for bipolar disorder. Biol Psychiatry 84: 803–809.

19. Mesbah R, Koenders MA, van der Wee NJ, Giltay EJ, van Hemert AM, de Leeuw M (2023): Association between the fronto-limbic network and cognitive and emotional functioning in individuals with bipolar disorder: a systematic review and meta- analysis. JAMA Psychiatry. Retrieved April 19, 2024, from https://jamanetwork.com/journals/jamapsychiatry/article-abstract/2802944

20. Hirschfeld RM, Williams JB, Spitzer RL, Calabrese JR, Flynn L, Keck Jr PE, et al. (2000): Development and validation of a screening instrument for bipolar spectrum disorder: the Mood Disorder Questionnaire. Am J Psychiatry 157: 1873–1875.

21. Saunders KEA, Cipriani A, Rendell J, Attenburrow M-J, Nelissen N, Bilderbeck AC, et al. (2016): Oxford Lithium Trial (OxLith) of the early affective, cognitive, neural and biochemical effects of lithium carbonate in bipolar disorder: study protocol for a randomised controlled trial. Trials 17: 116.

22. Rush AJ, Trivedi MH, Ibrahim HM, Carmody TJ, Arnow B, Klein DN, et al. (2003): The 16- Item Quick Inventory of Depressive Symptomatology (QIDS), clinician rating (QIDS-C), and self-report (QIDS-SR): a psychometric evaluation in patients with chronic major depression. Biol Psychiatry 54: 573–583.

23. Altman EG, Hedeker D, Peterson JL, Davis JM (1997): The Altman self-rating mania scale. Biol Psychiatry 42: 948–955.

24. Bürkner P-C, Charpentier E (2018): Monotonic effects: A principled approach for including ordinal predictors in regression models.

25. Watson D, Clark LA, Tellegen A (1988): Development and validation of brief measures of positive and negative affect: the PANAS scales. J Pers Soc Psychol 54: 1063.

26. R Core Team (2020): R: A Language and Environment for Statistical Computing. Vienna, Austria: R Foundation for Statistical Computing. Retrieved from https://www.R-project.org/

27. Carpenter B, Gelman A, Hoffman MD, Lee D, Goodrich B, Betancourt M, et al. (2017): Stan : A Probabilistic Programming Language. J Stat Softw 76. 10.18637/jss.v076.i01

28. Bürkner P-C (2017): Advanced Bayesian multilevel modeling with the R package brms. ArXiv Prepr ArXiv170511123.

29. Bürkner P-C (2017): brms: An R package for Bayesian multilevel models using Stan. J Stat Softw 80: 1–28.

30. Wickham H, François R, Henry L, Müller K (2021): Dplyr: A Grammar of Data Manipulation. Retrieved from https://CRAN.R-project.org/package=dplyr

31. Kassambara A (n.d.): ggpubr:‘ggplot2’Based Publication Ready Plots (2018). R Package Version 02.

32. Lüdecke D, Bartel A, Schwemmer C (2019): Package ‘sjPlot.’

33. Subirana I, Sanz H, Vila J (2014): Building bivariate tables: the compareGroups package for R. J Stat Softw 57: 1–16.

34. Lenth RV (2021): Emmeans: Estimated Marginal Means, Aka Least-Squares Means. Retrieved from https://CRAN.R-project.org/package=emmeans

35. Xiao N (2018): Ggsci: Scientific Journal and Sci-Fi Themed Color Palettes for “Ggplot2*.”* Retrieved from https://CRAN.R-project.org/package=ggsci

36. Morey RD, Hoekstra R, Rouder JN, Wagenmakers E-J (2016): Continued misinterpretation of confidence intervals: response to Miller and Ulrich. Psychon Bull Rev 23: 131–140.

37. Nassar MR, Frank MJ (2016): Taming the beast: extracting generalizable knowledge from computational models of cognition. Curr Opin Behav Sci 11: 49–54.

38. Scholl J, Klein-Flügge M (2018): Understanding psychiatric disorder by capturing ecologically relevant features of learning and decision-making. Behav Brain Res 355: 56–75.

39. Reinharth J, Braga R, Serper M (2017): Characterization of Risk-Taking in Adults With Bipolar Spectrum Disorders. J Nerv Ment Dis 205: 580.

40. Lasagna CA, Pleskac TJ, Burton CZ, McInnis MG, Taylor SF, Tso IF (2022): Mathematical Modeling of Risk-Taking in Bipolar Disorder: Evidence of Reduced Behavioral Consistency, With Altered Loss Aversion Specific to Those With History of Substance Use Disorder [no. 1]. Comput Psychiatry 6: 96–116.

41. Jenkinson M, Beckmann CF, Behrens TEJ, Woolrich MW, Smith SM (2012): FSL. NeuroImage 62: 782–790.

42. Beckmann CF, Jenkinson M, Smith SM (2003): General multilevel linear modeling for group analysis in FMRI. Neuroimage 20: 1052–1063.

43. Woolrich MW, Behrens TE, Beckmann CF, Jenkinson M, Smith SM (2004): Multilevel linear modelling for FMRI group analysis using Bayesian inference. Neuroimage 21: 1732–1747.

44. Chau BKH, Sallet J, Papageorgiou GK, Noonan MP, Bell AH, Walton ME, Rushworth MFS (2015): Contrasting Roles for Orbitofrontal Cortex and Amygdala in Credit Assignment and Learning in Macaques. Neuron 87: 1106–1118.

45. Panchal, P, Nelissen, N, McGowen, N, Atkinson, LZ, Saunders, KEA, Harrison, PJ, et al. (in submission): Identifying mood instability and circadian rest-activity patterns using digital remote monitoring and actigraphy in participants at risk for bipolar disorder.

46. Holmes EA, Bonsall MB, Hales SA, Mitchell H, Renner F, Blackwell SE, et al. (2016): Applications of time-series analysis to mood fluctuations in bipolar disorder to promote treatment innovation: a case series [no. 1]. Transl Psychiatry 6: e720–e720.

47. Tsanas A, Saunders KEA, Bilderbeck AC, Palmius N, Osipov M, Clifford GD, et al. (2016): Daily longitudinal self-monitoring of mood variability in bipolar disorder and borderline personality disorder. J Affect Disord 205: 225–233.

48. Rutledge RB, Skandali N, Dayan P, Dolan RJ (2014): A computational and neural model of momentary subjective well-being. Proc Natl Acad Sci 111: 12252–12257.

49. Vinckier F, Rigoux L, Oudiette D, Pessiglione M (2018): Neuro-computational account of how mood fluctuations arise and affect decision making [no. 1]. Nat Commun 9: 1708.

50. Alloy LB, Nusslock R, Boland EM (2015): The development and course of bipolar spectrum disorders: An integrated reward and circadian rhythm dysregulation model. Annu Rev Clin Psychol 11: 213–250.

51. Whitton AE, Treadway MT, Pizzagalli DA (2015): Reward processing dysfunction in major depression, bipolar disorder and schizophrenia. Curr Opin Psychiatry 28: 7–12.

52. Neville V, Dayan P, Gilchrist ID, Paul ES, Mendl M (2021): Dissecting the links between reward and loss, decision-making, and self-reported affect using a computational approach. PLOS Comput Biol 17: e1008555.

53. Hayden BY (2018): Economic choice: the foraging perspective. Curr Opin Behav Sci 24: 1– 6.

54. Lieder F, Griffiths TL (2020): Resource-rational analysis: Understanding human cognition as the optimal use of limited computational resources. Behav Brain Sci 43.

55. Oaksford M, Chater N (2020): New Paradigms in the Psychology of Reasoning. Annu Rev Psychol 71: 305–330.

56. Kolling N, Wittmann M, Rushworth MFS (2014): Multiple Neural Mechanisms of Decision Making and Their Competition under Changing Risk Pressure. Neuron 81: 1190– 1202.

57. O’Reilly RC, Hazy TE, Mollick J, Mackie P, Herd S (2014): Goal-driven cognition in the brain: a computational framework. ArXiv Prepr ArXiv14047591.

58. Korn CW, Bach DR (2015): Maintaining homeostasis by decision-making. PLoS Comput Biol 11: e1004301.

59. Juechems K, Summerfield C (2019): Where Does Value Come From? Trends Cogn Sci 23: 836–850.

60. Juechems K, Balaguer J, Castañón SH, Ruz M, O’Reilly JX, Summerfield C (2019): A network for computing value equilibrium in the human medial prefrontal cortex. Neuron 101: 977–987.

61. Lam D, Wong G, Sham P (2001): Prodromes, coping strategies and course of illness in bipolar affective disorder–a naturalistic study. Psychol Med 31: 1397–1402.

62. Eckblad M, Chapman LJ (1986): Development and validation of a scale for hypomanic personality. J Abnorm Psychol 95: 214.

63. Liu X, Hairston J, Schrier M, Fan J (2011): Common and distinct networks underlying reward valence and processing stages: A meta-analysis of functional neuroimaging studies. Neurosci Biobehav Rev 35: 1219–1236.

64. Rushworth MF, Kolling N, Sallet J, Mars RB (2012): Valuation and decision-making in frontal cortex: one or many serial or parallel systems? Curr Opin Neurobiol 22: 946– 955.

65. Sescousse G, Caldú X, Segura B, Dreher J-C (2013): Processing of primary and secondary rewards: A quantitative meta-analysis and review of human functional neuroimaging studies. Neurosci Biobehav Rev 37: 681–696.

66. Jauhar S, Fortea L, Solanes A, Albajes-Eizagirre A, McKenna PJ, Radua J (2021): Brain activations associated with anticipation and delivery of monetary reward: A systematic review and meta-analysis of fMRI studies. PLOS ONE 16: e0255292.

67. Fischer AG, Bourgeois-Gironde S, Ullsperger M (2017): Short-term reward experience biases inference despite dissociable neural correlates. Nat Commun 8: 1690.

68. Nusslock R, Young CB, Damme KSF (2014): Elevated reward-related neural activation as a unique biological marker of bipolar disorder: Assessment and treatment implications. Behav Res Ther 62: 74–87.

69. Volman I, Pringle A, Verhagen L, Browning M, Cowen PJ, Harmer CJ (2021): Lithium modulates striatal reward anticipation and prediction error coding in healthy volunteers. Neuropsychopharmacol Off Publ Am Coll Neuropsychopharmacol 46: 386–393.

70. Grunze H, Vieta E, Goodwin GM, Bowden C, Licht RW, Azorin J-M, et al. (2018): The World Federation of Societies of Biological Psychiatry (WFSBP) Guidelines for the Biological Treatment of Bipolar Disorders: Acute and long-term treatment of mixed states in bipolar disorder. World J Biol Psychiatry 19: 2–58.

## References

1. Pulcu E, Saunders KEA, Harmer CJ, Harrison PJ, Goodwin GM, Geddes JR, Browning M (2022): Using a generative model of affect to characterize affective variability and its response to treatment in bipolar disorder. Proceedings of the National Academy of Sciences 119: e2202983119.

2. Mars RB, Jbabdi S, Sallet J, O’Reilly JX, Croxson PL, Olivier E, et al. (2011): Diffusion-weighted imaging tractography-based parcellation of the human parietal cortex and comparison with human and macaque resting-state functional connectivity. Journal of Neuroscience 31: 4087–4100.

3. Neubert F-X, Mars RB, Sallet J, Rushworth MFS (2015): Connectivity reveals relationship of brain areas for reward-guided learning and decision making in human and monkey frontal cortex. PNAS 112: E2695–E2704.

4. Sallet J, Mars RB, Noonan MP, Neubert F-X, Jbabdi S, O’Reilly JX, et al. (2013): The Organization of Dorsal Frontal Cortex in Humans and Macaques. J Neurosci 33: 12255–12274.

5. Diedrichsen J, Balsters JH, Flavell J, Cussans E, Ramnani N (2009): A probabilistic MR atlas of the human cerebellum. NeuroImage 46: 39–46.

6. Neubert F-X, Mars RB, Thomas AG, Sallet J, Rushworth MF (2014): Comparison of human ventral frontal cortex areas for cognitive control and language with areas in monkey frontal cortex. Neuron 81: 700–713.

7. Panchal, P, Nelissen, N, McGowen, N, Atkinson, LZ, Saunders, KEA, Harrison, PJ, et al. (in submission): Identifying mood instability and circadian rest-activity patterns using digital remote monitoring and actigraphy in participants at risk for bipolar disorder.

8. Nilsson ME, Suryawanshi S, Gassmann-Mayer C, Dubrava S, McSorley P, Jiang K (2013): Columbia–suicide severity rating scale scoring and data analysis guide. CSSRS Scoring Version 2: 1–13.

9. First MB (2014): Structured clinical interview for the DSM (SCID). The encyclopedia of clinical psychology 1–6.

10. Patton JH, Stanford MS, Barratt ES (1995): Factor structure of the Barratt impulsiveness scale. Journal of clinical psychology 51: 768–774.

11. Espie CA, Kyle SD, Hames P, Gardani M, Fleming L, Cape J (2014): The Sleep Condition Indicator: a clinical screening tool to evaluate insomnia disorder. BMJ open 4: e004183.

12. Zanarini MC, Vujanovic AA, Parachini EA, Boulanger JL, Frankenburg FR, Hennen J (2003): A screening measure for BPD: The McLean screening instrument for borderline personality disorder (MSI-BPD). Journal of personality disorders 17: 568–573.

13. Oliver MN, Simons JS (2004): The affective lability scales: Development of a short-form measure. Personality and individual differences 37: 1279–1288.

14. Larsen RJ (1984): Theory and Measurement of Affect Intensity as an Individual Difference Characteristic (Temperament, Emotion, Arousal). University of Illinois at Urbana-Champaign.

15. Rush AJ, Trivedi MH, Ibrahim HM, Carmody TJ, Arnow B, Klein DN, et al. (2003): The 16-Item Quick Inventory of Depressive Symptomatology (QIDS), clinician rating (QIDS-C), and self-report (QIDS-SR): a psychometric evaluation in patients with chronic major depression. Biological psychiatry 54: 573–583.

16. Altman EG, Hedeker D, Peterson JL, Davis JM (1997): The Altman self-rating mania scale. Biological psychiatry 42: 948–955.

17. Spitzer RL, Kroenke K, Williams JB, Löwe B (2006): A brief measure for assessing generalized anxiety disorder: the GAD-7. Archives of internal medicine 166: 1092–1097.

18. Group TE (1990): EuroQol-a new facility for the measurement of health-related quality of life. Health policy 16: 199–208.

19. Goodday SM, Atkinson L, Goodwin G, Saunders K, South M, Mackay C, et al. (2020): The True Colours Remote Symptom Monitoring System: A Decade of Evolution. Journal of Medical Internet Research 22: e15188.

20. Tsanas A, Saunders KEA, Bilderbeck AC, Palmius N, Osipov M, Clifford GD, et al. (2016): Daily longitudinal self-monitoring of mood variability in bipolar disorder and borderline personality disorder. Journal of affective disorders 205: 225–233.

21. Kolling N, Scholl J, Chekroud A, Trier HA, Rushworth MF (2018): Prospection, perseverance, and insight in sequential behavior. Neuron 99: 1069–1082.

22. Scholl J, Trier HA, Rushworth MF, Kolling N (2022): The effect of apathy and compulsivity on planning and stopping in sequential decision-making. PLoS biology 20: e3001566.

23. Kucukelbir A, Ranganath R, Gelman A, Blei DM (2015): Automatic variational inference in Stan. arXiv preprint arXiv:150603431.

24. Yao Y, Vehtari A, Simpson D, Gelman A (2018): Yes, but did it work?: Evaluating variational inference. International Conference on Machine Learning 5581–5590.

25. Gelman A, Vehtari A, Simpson D, Margossian CC, Carpenter B, Yao Y, et al. (2020): Bayesian workflow. *arXiv preprint arXiv:201101808*.

26. Nilsson H, Rieskamp J, Wagenmakers E-J (2011): Hierarchical Bayesian parameter estimation for cumulative prospect theory. Journal of Mathematical Psychology 55: 84–93.

27. Symmonds M, Wright ND, Bach DR, Dolan RJ (2011): Deconstructing risk: Separable encoding of variance and skewness in the brain. Neuroimage 58: 1139–1149.

28. Vrieze SI (2012): Model selection and psychological theory: A discussion of the differences between the Akaike information criterion (AIC) and the Bayesian information criterion (BIC). Psychological Methods 17: 228–243.

29. Bürkner P-C (2017): brms: An R package for Bayesian multilevel models using Stan. Journal of Statistical Software 80: 1–28.

30. Carpenter B, Gelman A, Hoffman MD, Lee D, Goodrich B, Betancourt M, et al. (2017): Stan : A Probabilistic Programming Language. Journal of Statistical Software 76. 10.18637/jss.v076.i01

31. Michael Betancourt (2017): How the shape of a weakly informative prior affects inferences. Retrieved from https://mc-stan.org/users/documentation/case-studies/weakly_informative_shapes.html

32. Lenth RV (2021): Emmeans: Estimated Marginal Means, Aka Least-Squares Means. Retrieved from https://CRAN.R-project.org/package=emmeans

33. Bonsall MB, Wallace-Hadrill SMA, Geddes JR, Goodwin GM, Holmes EA (2012): Nonlinear time-series approaches in characterizing mood stability and mood instability in bipolar disorder. Proceedings of the Royal Society B: Biological Sciences 279: 916–924.

34. Holmes EA, Bonsall MB, Hales SA, Mitchell H, Renner F, Blackwell SE, et al. (2016): Applications of time-series analysis to mood fluctuations in bipolar disorder to promote treatment innovation: a case series [no. 1]. Transl Psychiatry 6: e720–e720.

35. Deichmann R, Gottfried JA, Hutton C, Turner R (2003): Optimized EPI for fMRI studies of the orbitofrontal cortex. NeuroImage 19: 430–441.

36. Smith SM, Jenkinson M, Woolrich MW, Beckmann CF, Behrens TE, Johansen-Berg H, et al. (2004): Advances in functional and structural MR image analysis and implementation as FSL. Neuroimage 23: S208–S219.

37. Smith SM (2002): Fast robust automated brain extraction. Human brain mapping 17: 143–155.

38. Andersson JL, Jenkinson M, Smith S (2007): Non-linear registration aka Spatial normalisation FMRIB Technial Report TR07JA2. FMRIB Analysis Group of the University of Oxford.

39. Jenkinson M, Bannister P, Brady M, Smith S (2002): Improved optimization for the robust and accurate linear registration and motion correction of brain images. Neuroimage 17: 825–841.

40. Woolrich MW, Ripley BD, Brady M, Smith SM (2001): Temporal autocorrelation in univariate linear modeling of FMRI data. Neuroimage 14: 1370–1386.

